# Transmission dynamics of SARS-CoV-2 in the hospital setting

**DOI:** 10.1101/2021.04.28.21256245

**Authors:** Yin Mo, David W. Eyre, Sheila F. Lumley, Timothy M. Walker, Robert H. Shaw, Denise O’Donnell, Lisa Butcher, Katie Jeffery, Christl A. Donnelly, Oxford COVID infection review team, Ben S. Cooper

**Author notes:** Full list of members in the Oxford COVID infection review team is provided in the supplementary material section 9. Corresponding author: Yin Mo.

## Abstract

**Background:** SARS-CoV-2 can spread efficiently in hospitals, but the transmission pathways amongst patients and healthcare workers are unclear.

**Methods:** We analysed data from four teaching hospitals in Oxfordshire, UK, from January to October 2020. Associations between infectious SARS-CoV-2 individuals and infection risk were quantified using logistic, generalised additive and linear mixed models. Cases were classified as community- or hospital-acquired using likely incubation periods.

**Results:** Nine-hundred and twenty of 66184 patients who were hospitalised during the study period had a positive SARS-CoV-2 PCR test within the same period (1%). Out of these, 571 patients had their first positive PCR tests while hospitalised (62%), and 97 of these occurred at least seven days after admission (11%). Amongst the 5596 healthcare workers, 615 (11%) tested positive during the study period using PCR or serological tests. For susceptible patients, one day in the same ward with another patient with hospital-acquired SARS-CoV-2 was associated with an additional eight infections per 1000 susceptible patients (95%CrI 6-10). Exposure to an infectious patient with community-acquired COVID-19 or to an infectious healthcare worker was associated with substantially lower infection risks (2/1000 susceptible patients/day, 95%CrI 1-2). As for healthcare worker infections, exposure to an infectious patient with hospital-acquired SARS-CoV-2 or to an infectious healthcare worker were both associated with an additional one infection per 1000 susceptible healthcare workers per day (95%CrI 1-2). Exposure to an infectious patient with community-acquired SARS-CoV-2 was associated with half this risk (0.5/1000 susceptible healthcare workers/day, 95%CrI 0.3-0.7).

**Interpretation:** Exposure to patients with hospital-acquired SARS-CoV-2 poses a substantial infection risk. Infection control measures to limit nosocomial transmission must be optimised to protect both staff and patients from SARS-CoV-2 infection.

**Funding:** National Institute for Health Research Health Protection Research Unit (NIHR HPRU) in Healthcare Associated Infections and Antimicrobial Resistance at Oxford University in partnership with Public Health England (PHE) (NIHR200915). Medical Research Council, Nosocomial transmission of SARS-CoV-2 (MR/V028456/1).

**Research in context:** *Evidence before this study:* We searched the PubMed database using the search terms (”COVID-19” OR ”SARS-CoV-2”) AND (”nosocomial” OR ”hospital”) AND (”transmission”) in either the abstracts or titles, for English-language articles published up to March 31, 2021. This returned 748 results, out of which ten reported transmission events in the hospital setting quantitatively. These publications can be broadly categorised to epidemiological descriptions of isolated outbreaks (5) or contact tracing of patients exposed to infected healthcare workers (1), retrospective cohort studies involving a particular group of patients, e.g., patients who underwent surgical procedures (2), and using genomic sequencing to identify transmission clusters (2). None of the studies reported the comparative transmission rates of SARS-CoV-2 amongst patients and staff.

*Added value of this study:* This study reports the analysis of a large observational dataset collected from a group of hospitals in the UK over eight months, consisting of both hospitalised patients and healthcare workers. Based on these detailed individual-level data, we quantified the associations between patient and healthcare worker characteristics and risks for acquiring nosocomial SARS-CoV-2 infection after adjusting for their exposures to SARS-CoV-2. Over the study period, we describe how risk of acquisition changes both with calendar time and over a patient’s hospital stay. By linking the presence of infected and susceptible patients and healthcare workers by time and ward locations, we quantify the relative importance of the transmission pathways for both the susceptible patients and healthcare workers.

*Implications of all the available evidence:* Nosocomial transmission of SARS-CoV-2 is common. Identifying the drivers of SARS-CoV-2 transmissions in the hospital setting is essential for designing infection prevention and control policies to minimise the added pressure from such events on our health systems. We found that newly infected patients who acquired SARS-CoV-2 in the hospital pose the highest risk of onward transmission to other patients and healthcare workers. Infection control and prevention efforts need to be enhanced around these patients to prevent further transmissions and studies assessing the effectiveness of these policies are needed.

## Introduction

Nosocomial transmission and outbreaks of SARS-CoV-2 have been frequently reported in various healthcare settings since the beginning of the pandemic. [1–6] Reported proportions of hospitalised COVID-19 patients suspected to have acquired SARS-CoV-2 in the hospitals vary widely, ranging from *<*1% to 20%, [7–10] and a national data linkage study in England estimated that 15% of laboratory-confirmed cases among hospital patients were healthcare-associated. [11]

Nosocomial transmission of SARS-CoV-2 is of considerable concern. Hospitalised patients are especially vulnerable to COVID-19 associated complications and mortality. [2] Infected patients who are asymptomatic or become symptomatic after discharge from the hospital may contribute to the further spread of SARSCoV-2 in the community and nursing homes. Healthcare workers (HCW) are disproportionately infected with SARS-CoV-2. [12–15] They may be a key source of viral transmission to patients and fellow colleagues. Reduced staff workforce due to SARS-CoV-2 infection may compromise the clinical management of patients and infection prevention and control measures. These threats remain relevant despite the introduction of vaccines as novel variants can reduce the protection afforded, and their efficacy preventing onward transmissions may only be partial.

Analysis of detailed individual-level data including information on patients at risk of becoming infected has been lacking and the relative importance of different transmission pathways (e.g. patient to HCW, HCW to patient, HCW to HCW and patient to patient) and has not, to our knowledge, previously been quantified. [16] Improved understanding of the drivers of nosocomial SARS-CoV2 infection is of potential value for improving targeting of infection prevention and control activities in hospitals.

The objectives of this analysis are to use high resolution individual-level data to quantify associations between patient characteristics and risks for acquiring nosocomial SARS-CoV-2 infection after adjusting for exposures, describe how risk of acquisition changes both with calendar time and over a patient’s hospital stay, and provide evidence about the relative importance of different transmission pathways for both patients and HCW.

## Methods

### Study cohort

Data were obtained from Oxford University Hospitals, a group of four teaching hospitals (denoted hospital A-D) in Oxfordshire, UK from 12 January 2020 to 2 October 2020. Two of the four hospital sites (hospitals A and C) have an Emergency Department, and admitted symptomatic SARS-CoV-2 patients directly from the community. Patient data included patient demographics, location in the hospital on every day of stay, total length of stay, and SARS-CoV-2 PCR test results (supplementary section 7 for details of PCR assays).

SARS-CoV-2 infections in hospital HCW were identified using PCR results from symptomatic and asymptomatic testing at the hospital. Symptomatic testing was offered to staff from 27 March 2020 onwards and staff could participate in a voluntary asymptomatic screening programme from 23 April 2020 onwards, offering testing up to once every two weeks. Additionally, probable infections prior to widespread availability of testing were identified in staff without a positive PCR result, but who were either anti-nucleocapsid or anti-spike IgG antibody positive and recalled a date of onset of symptoms consistent with COVID-19. These symptoms were the presence of fever and new persistent cough, or anosmia or loss of taste. [17, 18] Hospital HCW and patients who were on the same wards during the study period were included in the analysis.

Data were classified as time-fixed and time-varying variables. Time-fixed variables included age at admission, sex and ethnicity routinely collected in hospital records. Time-varying variables included patients’ ward and hospital location, and the number of other patients and HCW known to be infected (and likely infectious) present on the same ward while a patient was at risk of becoming infected with SARS-CoV-2.

Deidentified patient data and data from HCW testing were obtained from electronic healthcare records using the Infections in Oxfordshire Research Database (IORD) which has generic Research Ethics Committee, Health Research Authority and Confidentiality Advisory Group approvals (19/SC/0403, 19/CAG/0144).

### Definitions and assumptions

#### Incubation period

We assumed that each individual could only be infected once, and hence patients and HCW were no longer at risk for acquiring SARS-CoV-2 after their first positive PCR test. The day each patient with a potential nosocomial infection became infected is unknown, but based on knowledge of the incubation period distribution we expect it to be one to 20 days prior to the date of symptom onset, with 83% falling between 3-7 days. [19] For a given incubation period, *d*, we assume that each patient with a nosocomial infection became infected *d* days before the date of symptom onset.

Among 245 inpatients testing positive after developing SARS-CoV-2 symptoms during hospitalisation, the mean interval between symptom onset and their swab for PCR-testing was one day (interquartile range 1-3). Consequently, we assumed that swabs for SARS-CoV-2 PCR tests after hospital admission were taken in response to COVID-19-like symptom onset one day earlier or, in asymptomatic cases, the swabs were assumed to have been taken one day after the incubation period. The date of each patient’s first positive PCR test refers to the date the swab was obtained, rather than tested if this differed (figure 1).

**Figure 1:**
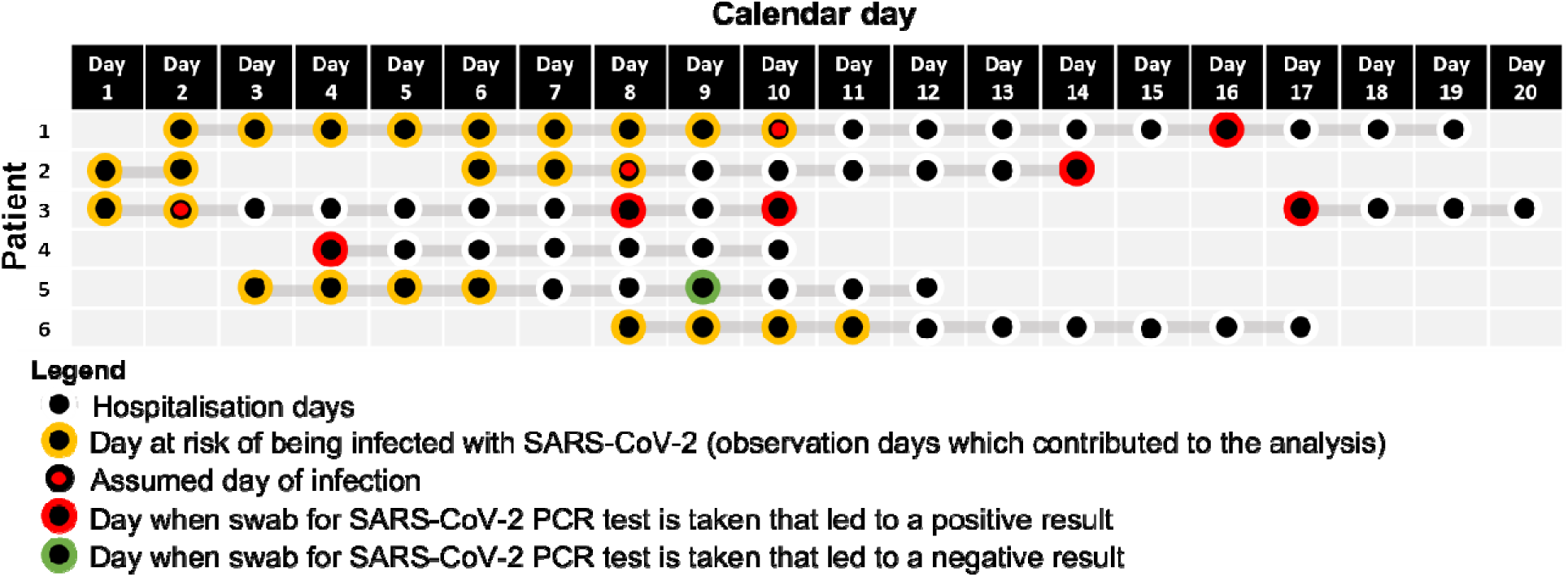
Illustration of assumed incubation periods and the data analysed for six example patients. We assumed that PCR tests were performed one day after developing symptoms consistent with COVID-19. In this schematic, an incubation period of five days was used. Each hospitalised patient day from admission until (and including) the day of the assumed infection event (i.e. six (incubation period plus one) days prior to the swab leading to the patient’s first positive PCR test) or six days prior to the day of discharge or death (whichever occurred first) was considered an observation where the patient was at risk of becoming infected. Each observation, unique to a specific patient on a specific day, therefore corresponds to an outcome six days later when the patient could potentially have a swab taken for a SARS-CoV-2 PCR test. An observation had a positive outcome (value of 1) if the patient had a positive PCR test for the first time resulting from a swab taken in the hospital six days later, and a negative outcome (value of 0) if the patient did not have a swab taken or had a swab taken resulting in a negative PCR test six days later. The risk factors, e.g., ward, number of infectious patients or healthcare workers in the same ward, for each observation were taken from the day of the observation itself when the corresponding patient was at risk of becoming infected.

### Definitions of nosocomial SARS-CoV-2 infections

Nosocomial SARS-CoV-2 infections have previously been defined as ‘probable’ when symptoms onset is on day 8-14 after admission and ‘definite’ when symptoms onset is on day >14 after admission. [20] These increasing thresholds correspond to higher certainties that a case is hospital-acquired (supplementary figure S5). [20] In this study, however, we used incubation periods that are the most likely to identify the exposure risk factors, i.e., the locations and infectious individuals the susceptible individuals were exposed to, which could have resulted in an observed infection event. Our baseline assumption was that the incubation period was five days (which is reported to be the median value [20]), and we therefore define hospital-acquired infections to be any PCR-confirmed SARS-CoV-2 infection where the patient was a hospital inpatient six days prior to the first positive PCR test. We also report results for sensitivity analyses assuming incubation periods of three and seven days. Community-acquired infections are defined to be any PCR-confirmed infections in patients who were not hospitalised in the 20 days prior to their first positive PCR tests.

### Accounting for varying infectiousness

We assumed that patients were infectious for a period of ten days starting a day after the day of presumed infection, consistent with estimates that 99.7% of onward infection takes place within the first ten days after the presumed infection event. [21, 26] HCW were assumed to be infectious from a day after the day of assumed infection to the day of symptom onset or one day prior to having a positive PCR test (i.e., staff were assumed to be absent from work after reporting symptoms consistent with SARS-CoV-2 infection or having a positive PCR test). In the main analyses presented in the Results section, we considered infectiousness to be binary. To account for time-varying infectiousness in relation to the time of presumed infection event, we repeated the analysis after scaling the numbers of infectious patients and HCW in a ward on a particular day by their relative infectiousness, using the generation time distribution derived by Ferretti *et al* [21] such that the sum of daily terms for a single infected patient or HCW who was present in the ward throughout their entire infectious period would equal one.

### Infection prevention and control measures

There were two major changes made to infection prevention and control measures during the study period. Prior to 1 April 2020 (phase 1) use of “level-1” personal protective equipment (PPE; apron, gloves, a surgical face mask and optional eye protection) was recommended for contact with patients known or suspected to have COVID-19 with use of “level-2” PPE (gown, gloves, eye protection, FFP3/N99 mask) for aerosol generating procedures. From 1 April 2020 (phase 2) universal level-1 PPE was used for all patients regardless of test results or clinical suspicion of COVID-19. From 25 April 2020 (phase 3), additionally, all patients were tested for SARS-CoV-2 by PCR on admission and at weekly intervals irrespective of symptoms. Further measures were implemented subsequently from June onwards, which include universal masking and social distancing amongst staff, contact tracing and isolation of exposed patients and HCW, establishment of COVID-19 dedicated areas, improved triage and recognition of atypical symptoms in elderly patients.

### Statistical models

We first performed exploratory analyses using univariable and multivariable logistic regression models to determine associations between risk factors and SARS-CoV-2 infection for given incubation periods (supplementary section 6 code block 1). To assess how well these individual demographic factors and infection pressures from infectious patients and healthcare workers on the same wards accounted for the nosocomial SARS-CoV-2 infections over calendar time, we used generalised additive models which allowed for the risk of infection to depend in a non-linear manner on the predictors (supplementary section 6 code block 2) The generalised additive models were implemented using the R package *mgcv*. [22]

We then modelled the patients’ and HCW’ daily risk of acquiring SARS-CoV-2 in the hospital using a generalised linear mixed model with an identity link (supplementary section 6 code block 3). This model allowed the daily probability of infection to scale linearly with infection pressure from HCW and patients and for their effects to be additive. These models were implemented with JAGS (version 4-10) which uses Markov chain Monte Carlo to generate a sequence of dependent samples from the posterior distribution of the parameters. [23]

Two versions of the models, one with interaction terms between the phases and forces of infection from patients and HCW, and one without the interaction terms, were performed. Model comparison was done using the Widely Applicable Information Criterion (WAIC) where lower values indicate improved model fit. [24]

All analysis was performed in R version 4.0.2. [25] The corresponding analysis code for the above models can be found in the supplementary material.

### Role of the funding source

The funders had no role in study design, data collection and analysis, decision to publish, or preparation of the manuscript. The views expressed in this publication are those of the authors and not necessarily those of the UK National Health Service, the National Institute for Health Research, the Department of Health or Public Health England, and other funders. All authors confirm that we had full access to all the data in the study and accept responsibility to submit for publication.

## Results

### Patient characteristics

There were 66,184 patients admitted to the four hospitals from 12 January to 2 October, 2020, a time period that covered only the first ‘wave’ of infection in the UK. Amongst these patients, 920 (920/ 66,184, 1%) had a positive SARS-CoV-2 PCR test. Out of these, 571 patients had their first positive PCR tests while hospitalised (62%), and 97 were on admission day seven or later (11%). The patient characteristics are shown in Table 1. The patients who likely acquired SARS-CoV-2 while in hospital (assuming incubation periods of 5, 3 or 7 days) were older, had longer lengths of stays and more readmissions compared to patients with no positive SARS-CoV-2 PCR tests.

**Table 1:** Characteristics of the patients included in the analysis.

| Patients testing positive for SARS-CoV-2<sup>#</sup><br>(n = 920) |  |  |  |  |  | Patients<br>did not<br>test<br>positive<br>for<br>SARS-CoV-2<sup>#</sup><br>(n =<br>65264) |
| --- | --- | --- | --- | --- | --- | --- |
|  |  | All<br>patients<br>tested<br>positive<br>(n = 920) | Hospital-acquired<br>infection<br>(assuming<br>an<br>incubation<br>period of 3<br>days)<br>(n = 133) | Hospital-acquired<br>infection<br>(assuming<br>an<br>incubation<br>period of 5<br>days)<br>(n = 130) | Hospital-acquired<br>infection<br>(assuming<br>an<br>incubation<br>period of 7<br>days)<br>(n = 120) |  |
| Age (mean age in<br>years, sd) |  | 67.9 (20.7) | 75.8 (17.3) | 76.9 (16.4) | 76.6 (16.6) | 49.1<br>(27.3) |
| Sex (n, %) | Female | 453 (49%) | 70 (53%) | 66 (51%) | 65 (54%) | 34887<br>(53%) |
|  | Male | 467 (51%) | 63 (47%) | 64 (49%) | 55 (46%) | 30350<br>(47%) |
| Ethnic groups (n,<br>%) | White | 630 (68%) | 107 (80%) | 105 (81%) | 100 (83%) | 46942<br>(72%) |
|  | Non-white | 111 (12%) | 0 (0%) | 2 (2%) | 2 (2%) | 5122<br>(8%) |
|  | Unknown | 179 (19%) | 26 (20%) | 23 (18%) | 18 (15%) | 13163<br>(20%) |
| Total<br>hospitalisation<br>days from<br>Jan to Oct 2020<br>(mean, sd) |  | 17.8 (22.2) | 38.6 (32.2) | 41.3 (32.5) | 42.1 (33.0) | 5.8 (11.8) |
| Total<br>number of<br>admissions<br>from Jan to<br>Oct 2020<br>(mean, sd) |  | 1.7 (1.2) | 1.9 (1.5) | 2.0 (1.5) | 2 (1.5) | 1.4 (1.2) |
| Admission<br>days to each<br>hospital from<br>Jan to Oct 2020<br>(n, %) | Hospital A | 855 (5%) | 248 (5%) | 279 (5%) | 284 (6%) | 57868<br>(15%) |
|  | Hospital B | 2846<br>(17%) | 959 (19%) | 1121 (21%) | 1076 (21%) | 37358<br>(10%) |
|  | Hospital C | 11417<br>(70%) | 3287 (64%) | 3238 (60%) | 3041 (60%) | 260247<br>(69%) |
|  | Hospital D | 1279 (8%) | 634 (12%) | 731 (14%) | 653 (13%) | 23226<br>(6%) |
| Number of<br>SARS-CoV-2 |  | 2.7 (2.7) | 3.5 (3.2) | 3.8 (3.4) | 3.8 (3.4) | 0.9 (1.7) |

| PCR tests per patient (mean, sd) |  |  |  |  |  |  |
| --- | --- | --- | --- | --- | --- | --- |
| Admission days to each ward type during infectious period<sup>§</sup> | General Ward | 3283 (87%) | 1234 (97%) | 1121 (97%) | 946 (97%) |  |
|  | ICU/ HDU* | 471 (13%) | 42 (3%) | 36 (3%) | 34 (3%) |  |
| Admission days to each ward type during at-risk period<sup>+</sup> | General Ward | 4737 (96%) | 1924 (95%) | 2254 (95%) | 2252 (95%) | 134001 (92%) |
|  | ICU/ HDU* | 178 (4%) | 100 (5%) | 125 (5%) | 122 (5%) | 11968 (8%) |
| At-risk days per patient (mean, sd) |  | 5.3 (11.5) | 15.2 (17.3) | 18.3 (18.1) | 19.8 (17.8) | 2.2 (10.3) |

<sup>#</sup> All patients included in the table had at least one day of inpatient stay during the observation period between 12 January and 2 October 2020.
\* ICU/ HDU: Intensive care units/ High-dependency units
<sup>§</sup> Infectious period: Patients were considered infectious from the day after infection to ten days after infection. [21]
<sup>+</sup> At-risk period: Patients were considered to be at risk of being infected with SARS-CoV-2 from admission to either discharge/ death or four, six or eight days before their first positive PCR tests, i.e., day of presumed infection.

Testing capacity increased substantially after the beginning of March 2020 (figure 2A). The weekly incidence of newly detected SARS-CoV-2 infections in the four hospitals, including both community-acquired and nosocomial cases, peaked between March and May 2020.

**Figure 2:**
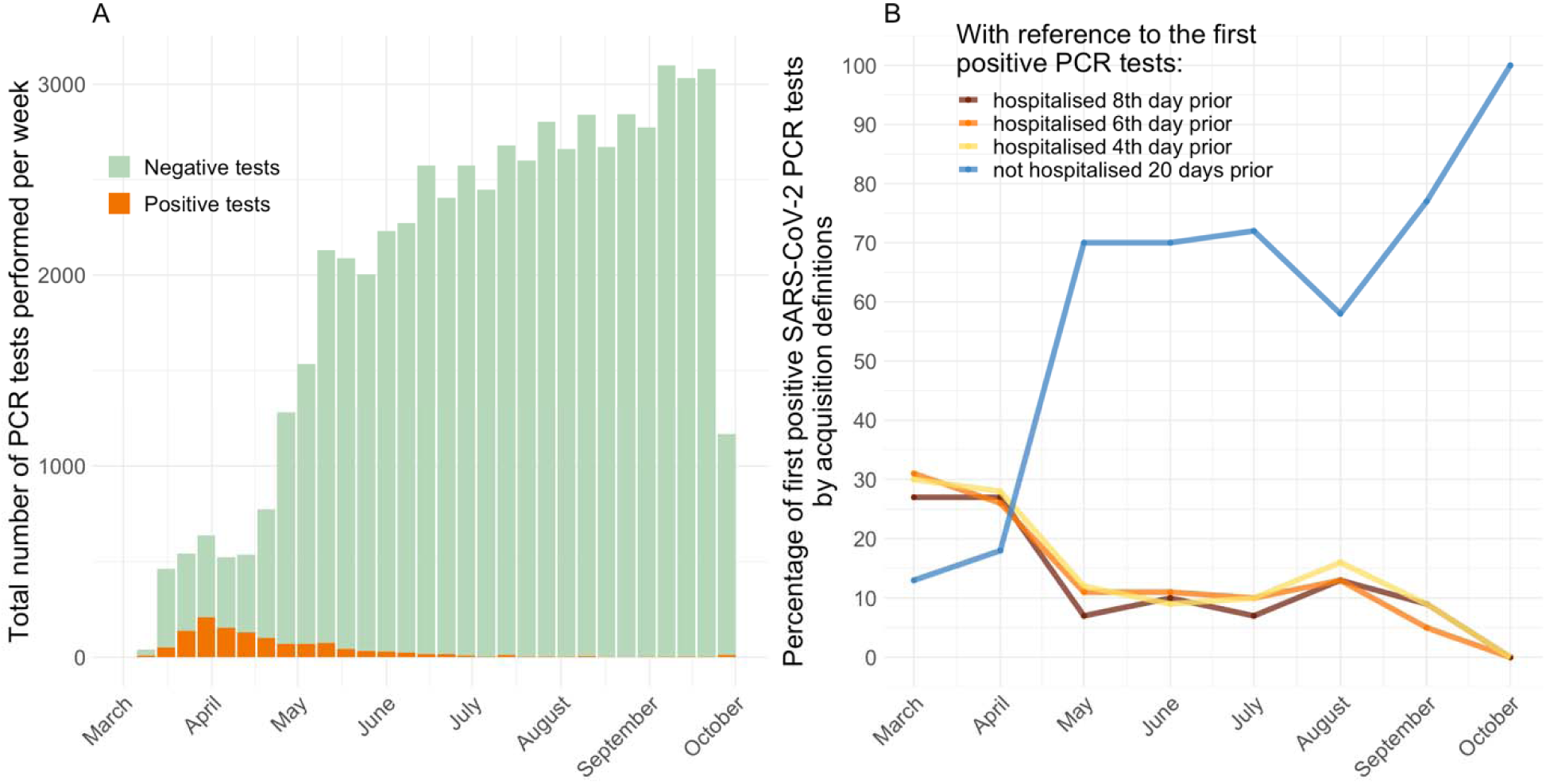
Weekly sums of SARS-CoV-2 PCR tests performed during March to October 2020 (Panel A). The stacked green bars indicate the number of negative tests. The stacked orange bars indicate the number of positive tests. Percentage of first positive SARS-CoV-2 PCR tests classified by different types of acquisition (Panel B). The colours represent patients who were inpatients on the eighth (red), sixth (orange), and forth day (yellow) prior to their first positive tests, and who were not hospitalised in the 20 days prior to their first positive tests (blue). These classifications are not mutually exclusive, e.g., a patient who was admitted for ten days continuously prior to the first positive PCR test would contribute to all first three groups.

Two-hundred and seventy-one patients had at least one day of hospitalisation in the 20 days prior to being tested positive for SARS-CoV-2. Out of these patients, 130 (48%) were inpatients on their day of infection, based on an assumed incubation period of five days. One-hundred and two out of the 130 patients had at least one negative PCR test during day 1-5 of their hospitalisation (79%). The median length of stay for the admissions during which the patients were infected with SARS-CoV-2 was 21 days (interquartile range 13 to 35 days). The median day of hospitalisation when these patients were assumed to have been infected was day 8 (interquartile range 3 to 18 days).

### Healthcare worker characteristics

Out of a total of 13,514 HCW in the four hospitals participating in HCW testing at some time, 5,596 worked on a single ward only such that their SARS-CoV-2 status could be considered with patients admitted to the same wards in the analysis. Eleven percent (615/5596) were positive for SARS-CoV-2 during the study period. Amongst those who were positive, 57% (353/615) had a positive PCR test, while the rest were diagnosed based on serology.

The timelines of the numbers of susceptible patients and infectious patients and HCW on each ward showed that most of the peaks in the number of potential transmission events occurred between March and June 2020 (figure S1). On most wards there was considerable overlap between the time series for infectious HCW and patients and the time series of transmission events.

### Transmission risk to patients

We first used multivariable logistic regression to identify the factors associated with nosocomial transmission of SARS-CoV-2 to susceptible patients (table 3). Infection risk reduced with the introduction of more stringent infection prevention and control measures in phase three (adjusted odds ratio, aOR 0.25, 95%CI 0.14, 0.42). Presence of patients with hospital-acquired SARS-CoV-2 was associated with the highest risk of acquisition in susceptible patients (aOR, 1.76, 95%CI 1.51, 2.04), followed by the presence of infected HCW (aOR 1.45, 95%CI 1.22,1.71). The evidence that patients with community onset COVID-19 were associated with increased transmission was weaker (aOR 1.12, 95%CI 0.96,1.26).

**Table 2:** Characteristics of the healthcare workers included in the analysis.

|  |  | Positive for SARS-CoV-2<br>n = 615 | Negative for SARS-CoV-2<br>n = 4981 |
| --- | --- | --- | --- |
| Age (mean age in years, sd) |  | 39.5 (11.1) | 39.6 (11.7) |
| Sex (n, %) | Female | 485 (79%) | 3902 (78%) |
|  | Male | 130 (21%) | 1079 (22%) |
| Roles (n, %) | Doctor | 98 (16%) | 955 (19%) |
|  | Nurses | 306 (50%) | 1984 (40%) |
|  | Allied health | 136 (22%) | 1274 (26%) |
|  | Non-clinical staff | 75 (12%) | 768 (15%) |
| Hospital worked in during the observation period (n, %) | Hospital A | 97 (16%) | 972 (20%) |
|  | Hospital B | 91 (15%) | 454 (9%) |
|  | Hospital C | 379 (62%) | 3276 (66%) |
|  | Hospital D | 48 (8%) | 279 (6%) |
| Ward type worked in during the observation period (n, %) | General Ward | 569 (93%) | 4384 (88%) |
|  | ICU/ HDU* | 46 (7%) | 597 (12%) |

\* ICU/ HDU: Intensive care units/ High-dependency units

**Table 3:**
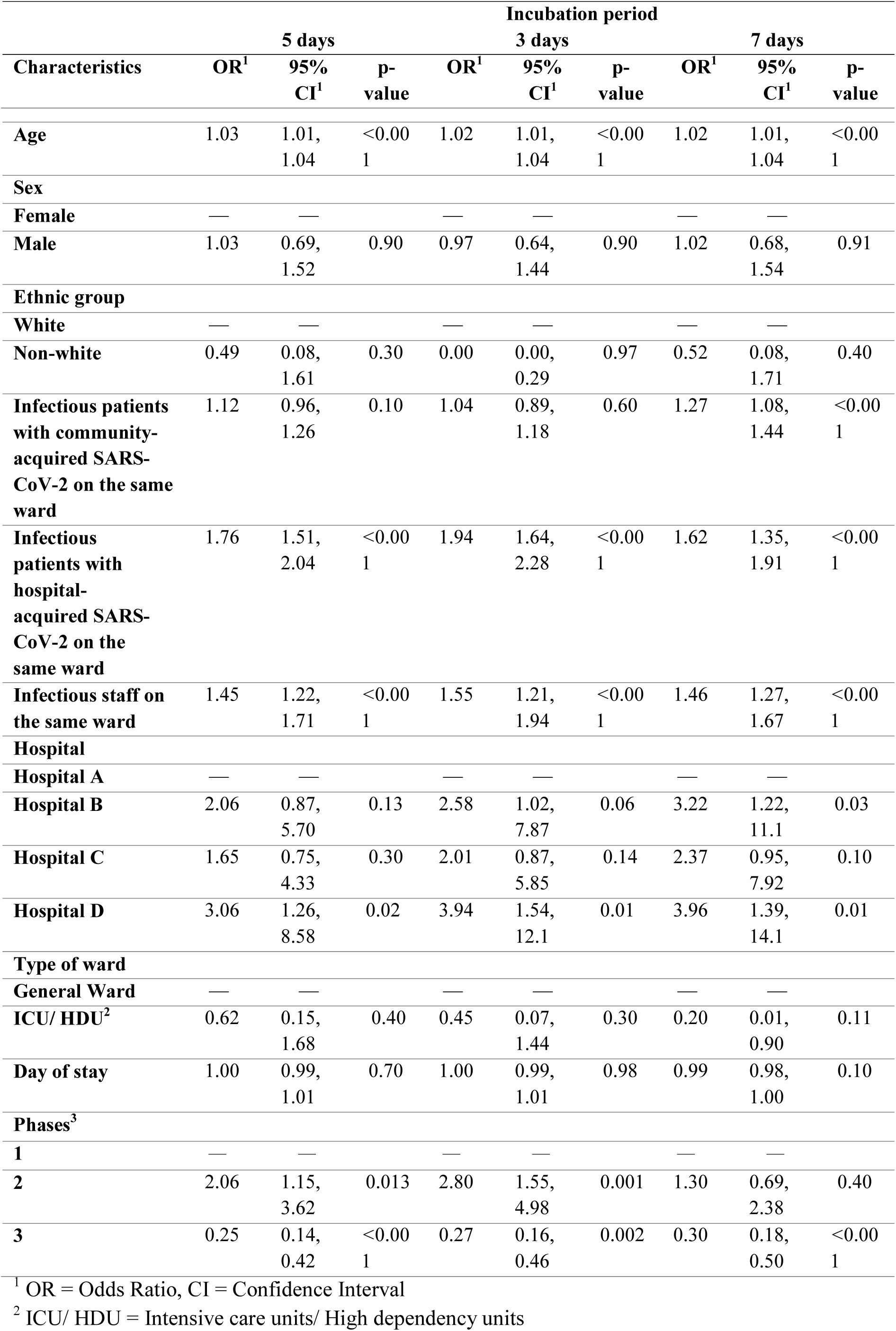
Predictors of SARS-CoV-2 infection in admitted patients during their hospital stay from multivariable logistic regression results.

To further investigate if the demographic variables and transmissions from infectious patients and HCW adequately accounted for patient acquisition of SARS-CoV-2, we used these variables in a generalised additive model (supplementary section 3.1 model *P*2). After adjusting for these variables, the results showed that the variation in the risk of nosocomial infection over the study period remained though at a reduced level, suggesting that transmission risks were incompletely accounted for (figure 3 panel A). We further used the above generalised additive model to explore how risk of nosocomial SARS-CoV-2 infection varied with day of hospitalisation (supplementary figure S2). This risk remained largely constant throughout a patient’s hospital stay once the numbers of infectious patients and HCW in the same ward were accounted for.

**Figure 3.**
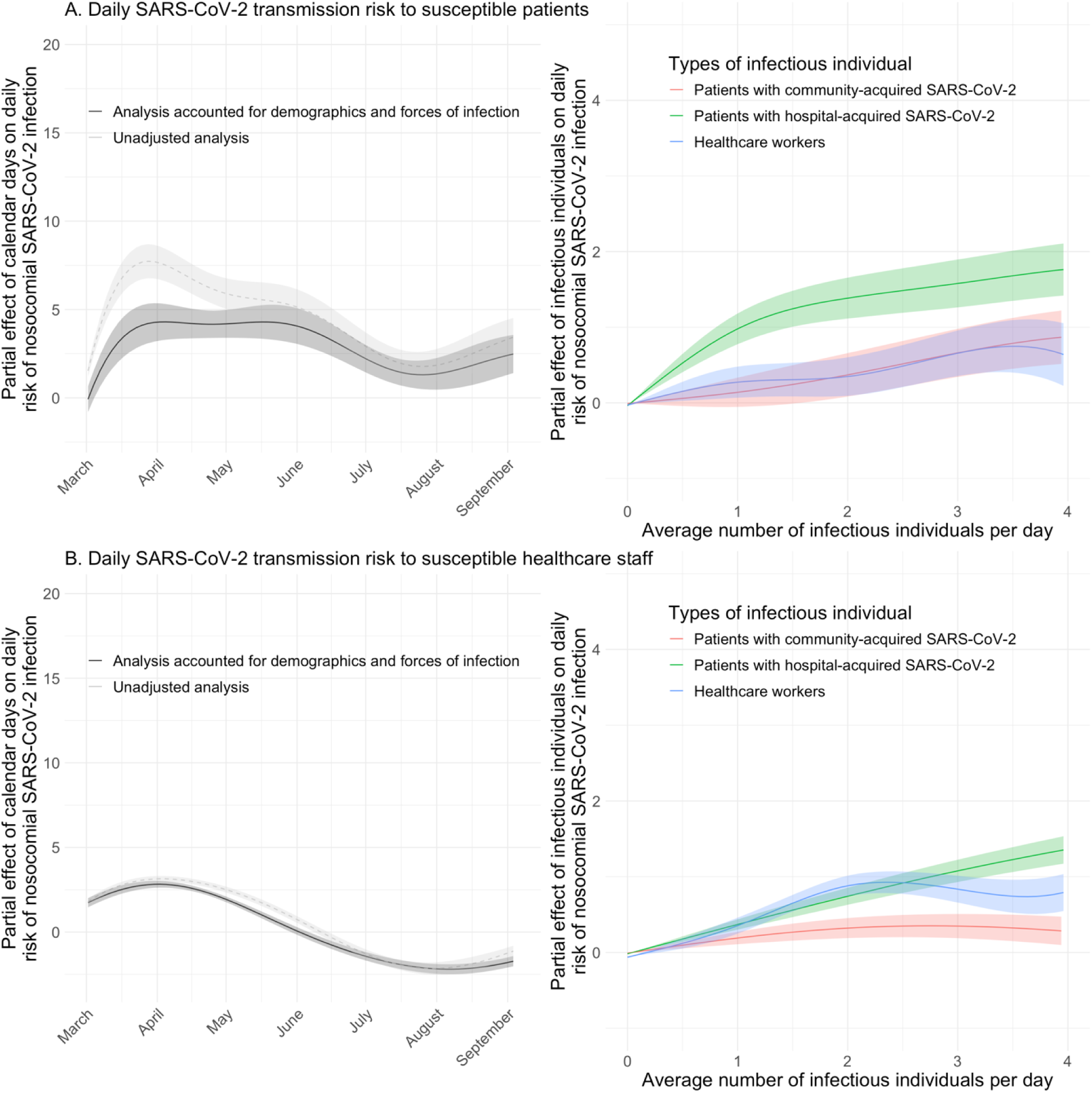
Daily transmission risk to susceptible patients (Panel A) and healthcare workers (Panel B) using a generalised additive model with a logit link. The smooth, non-linear partial effects of calendar day, infectious patients and healthcare workers on the daily risk of nosocomial SARS-CoV-2 infection are presented as coloured lines. These partial effects are the isolated effects of each group of infectious individuals on the binary outcome of assumed acquisition (yes/no) on each day as the dependent variable. Infectious patients and healthcare workers were both associated with increased risk of nosocomial infection. The presence of more infectious patients or healthcare workers in a ward on a given day was associated with higher transmission risk.

A shortcoming of the logistic regression model is that it assumed the effect of each additional infectious patient or HCW as multiplicative. Therefore, we used an alternative statistical model that allows each extra infectious individual to increase the probability of infection in an additive way (a generalised mixed model with an identity link). Infectious patients on the same ward who were assumed to have hospital-acquired SARS-CoV-2 showed the strongest association with acquisition of nosocomial COVID-19 in susceptible patients (figure 4). Using an assumed incubation period of 5 days, the absolute risk of acquiring SARS-CoV-2 per day of exposure to a patient with hospital-acquired SARS-CoV-2 infection was 0.75% (95% credible interval, CrI 0.55-0.95%). The risks of acquiring SARS-CoV-2 per day of exposure to an infectious patient who acquired SARS-CoV-2 in the community or to an infectious HCW were smaller. One day exposure to an infected HCW or patient with community-acquired COVID19 was associated with absolute risks of 0.20% (95%CrI 0.16-0.22%) and 0.17% (95%CrI 0.13-0.22%) respectively for susceptible patients.

**Figure 4:**
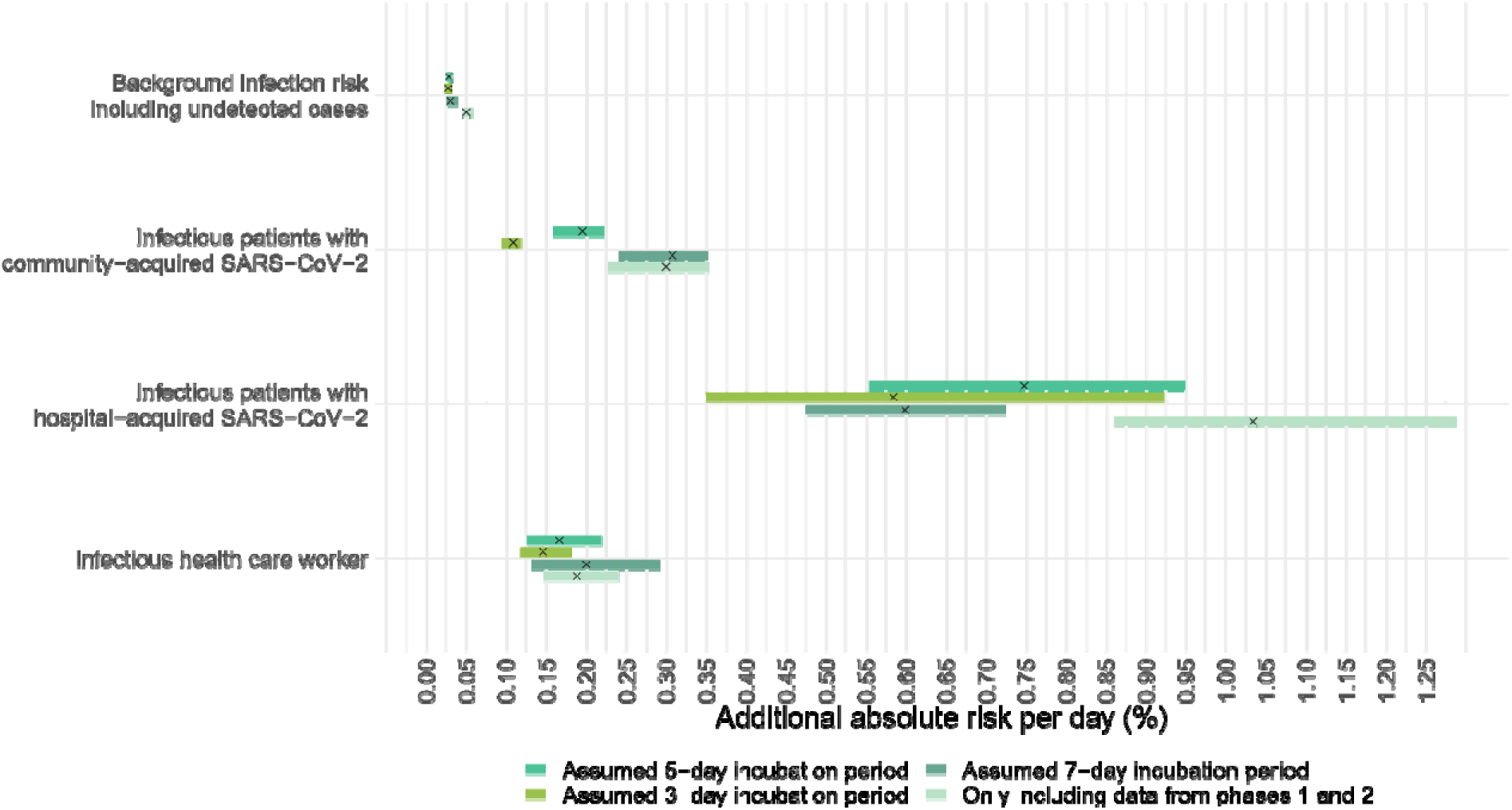
Additional risk of suspected nosocomial acquisition of SARS-CoV-2 experienced by a single susceptible patient contributed by i) infectious patients who acquired SARS-CoV-2 in the community (second row); ii) infectious patients who acquired SARS-CoV-2 in the hospital (third row) and iii) infectious healthcare workers (last row). A generalised mixed model with an identity link was used, with assumed nosocomial acquisition (yes/no) on each day as the dependent variable. Both the intercepts and slopes were allowed to vary by ward. The top row shows the variation of the intercepts of the model, which represent the background infection risk posed by infectious patients and healthcare workers who are undetected. Each horizontal bar represents the 95% credible interval of the estimate.

### Transmission risk to healthcare workers

We performed similar analyses to quantify the risk of transmission to HCW. The multivariable logistic regression results showed that nurses were at the highest risk of being infected with SARS-CoV-2 (aOR 1.58, 95%CI 1.18, 2.07). Working in the intensive-care or high-dependency units was protective against transmission (aOR 0.55, 95%CI 0.39, 0.75). Transmission risk reduced in phase three (aOR 0.43, 95%CI 0.34, 0.53). The number of infectious HCW and patients who had hospital-acquired SARS-CoV-2 on the same ward had the strongest associations with transmission to HCW (aOR 1.66, 95%CI 1.55,1.78 and aOR 1.45, 95%CI 1.32,1.58 respectively) (table 4).

**Table 4:** Predictors of SARS-CoV-2 infection in healthcare workers from multivariable logistic regression.

| Characteristics | Incubation period |  |  |  |  |  |  |  |  |
| --- | --- | --- | --- | --- | --- | --- | --- | --- | --- |
|  | 5 days |  |  | 3 days |  |  | 7 days |  |  |
|  | OR<sup>1</sup> | 95% CI<sup>1</sup> | p-value | OR<sup>1</sup> | 95% CI<sup>1</sup> | p-value | OR<sup>1</sup> | 95% CI<sup>1</sup> | p-value |
| Age<sup>2</sup> | 1.00 | 0.99, 1.01 | 0.80 | 1.00 | 0.99, 1.01 | 0.8 | 1.00 | 0.99, 1.01 | 0.92 |
| **Sex** |  |  |  |  |  |  |  |  |  |
| Female | — | — |  | — | — |  | — | — |  |
| Male | 1.23 | 0.97, 1.56 | 0.08 | 1.19 | 0.93, 1.50 | 0.2 | 1.19 | 0.93, 1.50 | 0.2 |
| **Role** |  |  |  |  |  |  |  |  |  |
| Doctor | — | — |  | — | — |  | — | — |  |
| Nurse | 1.58 | 1.18, 2.07 | 0.002 | 1.66 | 1.26, 2.21 | <0.001 | 1.50 | 1.15, 1.98 | 0.004 |
| Allied health | 1.03 | 0.76, 1.40 | 0.9 | 1.06 | 0.78, 1.45 | 0.7 | 0.93 | 0.68, 1.25 | 0.6 |
| Non-clinical staff | 1.03 | 0.72, 1.46 | 0.9 | 1.10 | 0.77, 1.56 | 0.6 | 0.94 | 0.66, 1.33 | 0.7 |
| Infectious patients with community-acquired SARS-CoV-2 on the same ward | 1.02 | 0.96, 1.09 | 0.5 | 1.04 | 0.98, 1.09 | 0.2 | 1.00 | 0.90, 1.10 | >0.9 |
| Infectious patients with hospital-acquired SARS-CoV-2 on the same ward | 1.45 | 1.32, 1.58 | <0.001 | 1.61 | 1.46, 1.76 | <0.001 | 1.44 | 1.32, 1.57 | <0.001 |
| Infectious staff on the same ward | 1.66 | 1.55, 1.78 | <0.001 | 1.83 | 1.65, 2.02 | <0.001 | 1.56 | 1.48, 1.64 | <0.001 |
| **Hospital** |  |  |  |  |  |  |  |  |  |
| Hospital A | — | — |  | — | — |  | — | — |  |
| Hospital B | 1.54 | 1.11, 2.13 | 0.01 | 1.66 | 1.21, 2.28 | 0.002 | 1.55 | 1.13, 2.13 | 0.007 |
| Hospital C | 1.18 | 0.92, 1.51 | 0.2 | 1.10 | 0.86, 1.41 | 0.5 | 1.16 | 0.91, 1.48 | 0.2 |
| Hospital D | 1.34 | 0.88, 2.00 | 0.2 | 1.42 | 0.94, 2.10 | 0.08 | 1.22 | 0.79, 1.83 | 0.4 |
| **Type of ward** |  |  |  |  |  |  |  |  |  |
| **General Ward** | — | — |  | — | — |  | — | — |  |
| **ICU/ HDU<sup>3</sup>** | 0.55 | 0.39, 0.75 | <0.001 | 0.57 | 0.41, 0.78 | 0.004 | 0.55 | 0.39, 0.75 | <0.001 |
| **Phase<sup>4</sup>** |  |  |  |  |  |  |  |  |  |
| **1** | — | — |  | — | — |  | — | — |  |
| **2** | 1.60 | 1.24, 2.05 | <0.001 | 1.67 | 1.30, 2.16 | <0.001 | 1.25 | 0.96, 1.61 | 0.09 |
| **3** | 0.43 | 0.34, 0.53 | <0.001 | 0.44 | 0.36, 0.55 | <0.001 | 0.39 | 0.32, 0.49 | <0.001 |

<sup>1</sup> OR = Odds Ratio, CI = Confidence Interval
<sup>2</sup> Age measured in years.
<sup>3</sup> ICU/ HDU = Intensive care units/ High dependency units
<sup>4</sup> In addition to phases, calendar days was included as a non-linear independent variable in the logistic regression, fitted with a linear spline function with two knots.

Using the alternative additive statistical model (figure 5), the strongest association was with other infectious staff and patients with hospital-acquired SARSCoV-2. However, the additional risks posed by exposures to these infectious HCW and patients to the susceptible HCW were less compared to the that for susceptible patients. A single day of exposure to infected HCW and patients with hospital-acquired SARS-CoV-2 patients on the same ward was associated with an increased absolute daily risk of 0.10% (95%CrI 0.04-0.20%). The corresponding increase in absolute daily risk from a day of exposure to an infected patient with community-acquired SARS-CoV-2 was 0.05% (95%CrI 0.03-0.07%).

**Figure 5:**
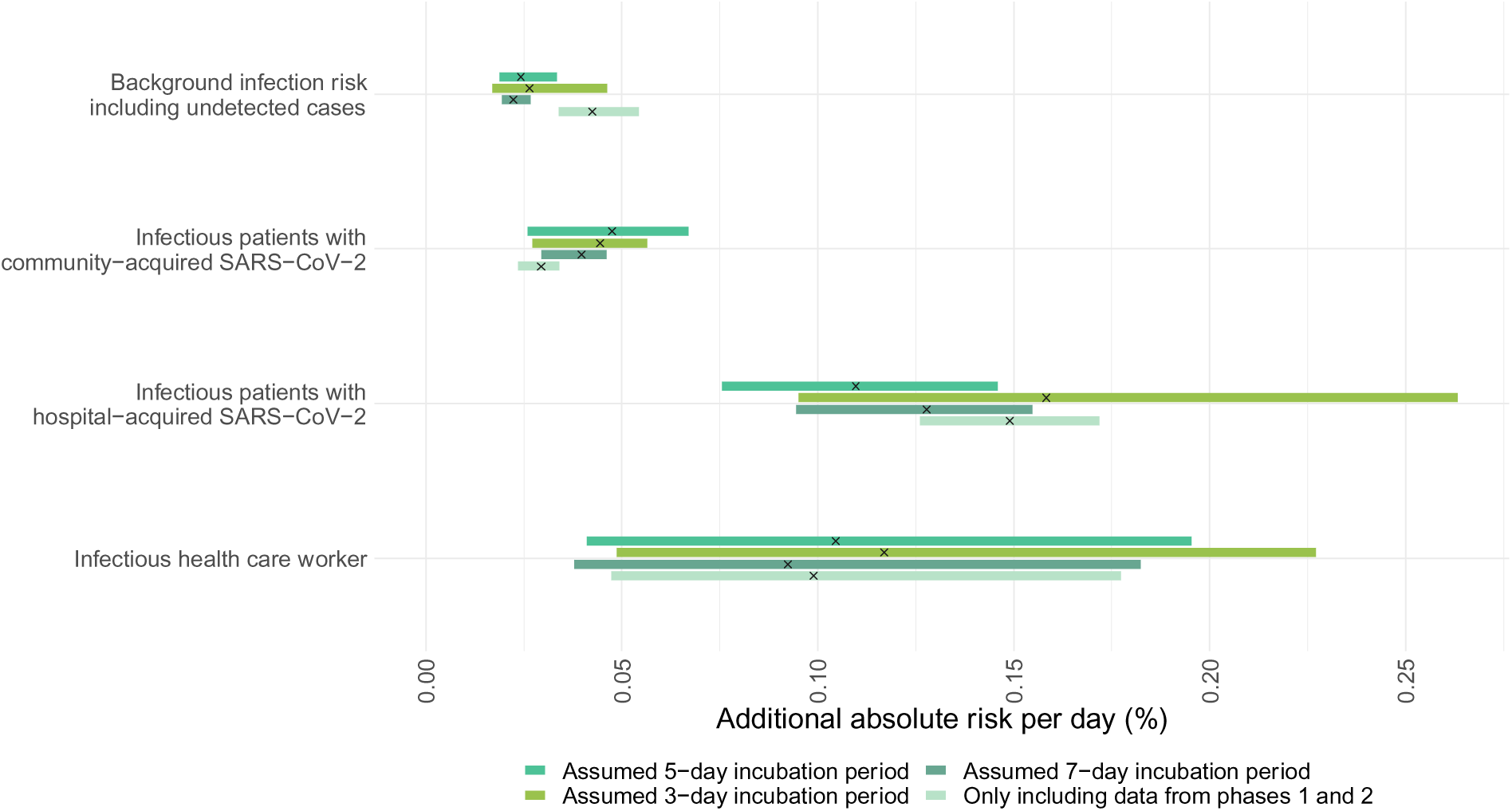
Additional risk of suspected nosocomial acquisition of SARS-CoV-2 experienced by a single susceptible healthcare worker contributed by i) infectious patients who acquired SARS-CoV-2 in the community (second row); ii) infectious patients who acquired SARS-CoV-2 in the hospital (third row) and iii) infectious healthcare workers (last row). A generalised mixed model with an identity link was used, with assumed nosocomial acquisition (yes/no) on each day as the dependent variable. Both the intercepts and slopes were allowed to vary by ward. The top row shows the variation of the intercepts of the model, which represent the background infection risk posed by infectious patients and healthcare workers who are undetected. Each horizontal bar represents the 95% credible interval of the estimate.

The background transmission risks to HCW including that from community sources and undetected cases amongst both HCW and patients were similar to those observed in the patients. The contribution of these undetected cases to the daily risk of SARS-CoV-2 acquisition was about 0.03% (95CrI 0.02-0.03%) and 0.02% (95%CrI 0.02-0.03%) for the susceptible patients and HCW respectively. Findings from sensitivity analyses which excluded data from phase three and using different prior distributions gave similar results as the main analyses (supplementary material section 5)

## Discussion

The consistent finding in the above analysis is that the patients who acquired SARS-CoV-2 in the hospital and, to a lesser degree, infectious healthcare workers likely working prior to the onset of symptoms, were the most strongly associated with increased risk of SARS-CoV-2 transmission in the hospital setting. In contrast, exposure to patients who had acquired SARS-CoV-2 in the community appeared to be associated with, at most, modest increases in the daily risk of infection for both healthcare staff and the other patients. We found evidence of a dose-response effect: exposure to more infectious patients and healthcare staff were both associated with increasing daily risk of acquiring SARS-CoV-2. These findings can parsimoniously be explained by newly infected individuals having high transmission potential to patients and staff. Multiple lines of evidence indicate that a substantial proportion of transmission precedes symptom onset and point to rapidly declining infectiousness with time since symptom onset. [21, 26] Secondly, patients who acquired SARS-CoV-2 in the community are more likely to first present with symptoms compatible with COVID-19 upon admission and be rapidly segregated from the susceptible population with careful implementation of infection prevention and control guidelines.

There are several limitations in our analysis. Firstly, the dates on which the infected patients first developed symptoms were not available. Hence, we needed to assume that the PCR test swabs were taken on the symptom onset dates. While this assumption is reasonable based on the analysis of a subset of data early in the pandemic, it is not true from phase three onwards when weekly screening of patients regardless of symptoms was implemented. We addressed this by performing sensitivity analysis comparing model outputs when using data collected during phase one and two versus phase three (supplementary material section 5). Secondly, we assumed that HCW were absent from work after the dates on which their first positive PCR test swabs were taken or COVID-19 symptoms were first self-reported. However, where HCW experienced minimal or no symptoms they may have continued to work. These issues could be further explored using HCW absentee data in subsequent analysis.

A key challenge in this analysis is that the times of infection are unknown. This has led to the adoption of various arbitrary cut-offs on length of stay prior to infection to define nosocomial infection. Further analysis using data augmentation methods may potentially overcome this to produce estimates that better account for different sources of uncertainty.

Other drivers of SARS-CoV-2 transmissions in the hospital setting not fully explained by infection pressures, which we did not capture in the analysis, may include variation in ward occupancy, community-acquired cases who did not develop symptoms until after hospitalisation, change in nature or frequency of SARS-CoV-2 exposures throughout hospitalisation, or could reflect frailties, i.e., those patients who have stayed 20 days and not been infected may be at lower risk of infection. However, recent work using detailed epidemiological and genomic data to infer transmission networks echoed our main finding that patients are more likely to be infected by other patients than by HCWs. [27]

Our findings support enhanced infection prevention and control efforts to prevent and identify early hospital-onset SARS-CoV-2 infection. Where either community or local ward prevalence is sufficiently high and resources permit, regular screening and prompt testing and identification of such patients is important. Similarly, measures to ensure symptomatic staff are not at work, including adequate sick pay arrangements, are vital. Regular staff screening is also likely to reduce transmission. Staff acquisition and transient asymptomatic carriage, contamination of equipment and the general environment or the air are possible mediators of transmission events assigned in the analysis as patient-to-patient and need further investigation. The relatively low risk of transmission associated with patients with suspected community-acquired COVID-19 suggests that for these patients the peak of their infectivity may have passed such that existing infection prevention and control policies including universal PPE, prompt testing and isolation of suspected or known cases [16] are sufficient to mitigate most of the remaining infectiousness. Our analysis shows that despite these measures patients and staff are at risk from newly infected individuals. Due to the difficulties in disentangling the effect of infection prevention and control measures and surges in SARS-CoV-2 in the community setting, we cannot provide conclusive evidence on how interventions around hospital-onset cases could be enhanced. However, others have suggested enhanced PPE for HCW and ventilation may play a role. [28–31]

In conclusion, our data provide strong evidence that newly infected patients pose a high risk of onward transmission to patients and healthcare workers in hospital. Further investigation is needed into how best to enhance infection control and prevention efforts around these patients.

## Data Availability

The datasets analysed during the current study are not publicly available as they contain personal data but are available from the Infections in Oxfordshire Research Database (https://oxfordbrc.nihr.ac.uk/research-themes-overview/antimicrobial-resistance-and-modernising-microbiology/infections-in-oxfordshire-research-database-iord/), subject to an application and research proposal meeting the ethical and governance requirements of the Database. All analysis codes are available at https://github.com/moyinNUHS/covid_HospTransmissionDynamics.

## Authors’ contributions

DWE and BSC conceptualized this work. MY, DWE and BSC performed the statistical analysis. MY drafted the first version of the manuscript. DWE, MY and KJ verified the underlying data. All authors reviewed and edited subsequent versions of the manuscript.

## Sources of funding

MY is supported by the Singapore National Medical Research Council Research Fellowship (Grant ref: NMRC/Fellowship/0051/2017). BSC acknowledges support from the Medical Research Council (Grant Ref: MR/V028456/1). TMW is a Wellcome Trust Clinical Career Development Fellow (214560/Z/18/Z). This work was also supported by the National Institute for Health Research Health Protection Research Unit (NIHR HPRU) in Healthcare Associated Infections and Antimicrobial Resistance at Oxford University in partnership with Public Health England (PHE) (NIHR200915), the NIHR Biomedical Research Centre, Oxford, and the NIHR HPRU in Emerging and Zoonotic Infections at University of Liverpool in partnership with PHE, in collaboration with Liverpool School of Tropical Medicine and the University of Oxford (NIHR200907).

## Conflicts of interest

DWE declares personal fees from Gilead outside the submitted work.

## Acknowledgement

The authors acknowledge valuable contributions from Omar Risk, Hannah Chase, Ishta Sharma, Sarah Peters, Tamsin Cargill, Grace Barnes, Josh Hamblin, Jenny Tempest-Mitchell, Archie Lodge, Sai Parepalli, Raghav Sudarshan, Hannah Callaghan, Imogen Vorley, Ashley Elder, Danica Fernandes, Gurleen Kaur, Bara’a Elhag, Edward David, Rumbi Mutenga, Dylan Riley, Emel Yildirim, and Naomi Hudson from Oxford University Hospitals NHS Foundation Trust and University of Oxford Medical School for data collection.

## Supplementary material

### 1. The timelines of potential nosocomial transmission events and the numbers of infectious patients and HCW.

**Figure S1.**
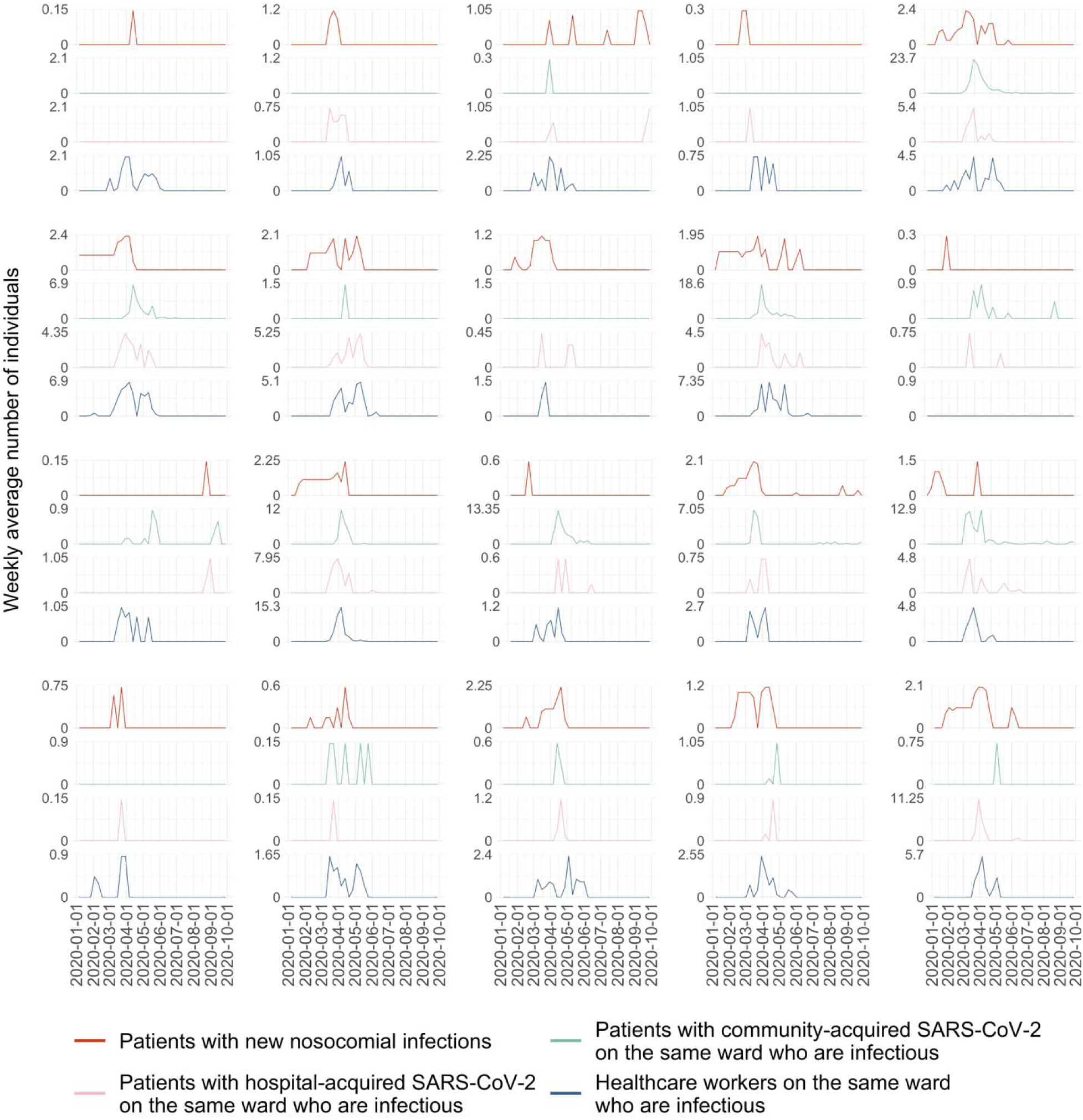
Weekly average numbers of transmission events and infectious patients and healthcare workers in wards with at least 30 members of staff tested and 20 available patient beds. The top row for each panel of graphs shows the weekly average number of patients who had a positive SARS-CoV-2 PCR test and who were defined to have been infected in the hospital on that ward during the indicated week based on the assumed incubation period of five days. The second and third rows for each panel show the numbers of infectious patients defined as having community-acquired SARS-CoV-2 infections (i.e., no hospitalisation in the 20 days prior to first positive tests), and patients who acquired SARS-CoV-2 in the hospital (i.e., inpatient on the sixth day prior to first positive tests) respectively. For these plots, patients were considered to be infectious for a period of ten days, starting one day after the day of the presumed infection event. [20] The last row shows the number of infectious healthcare workers. Healthcare workers were considered infectious from the day of the infection event until a day before their first positive PCR test or report of COVID-19 related symptoms, whichever was earlier.

### 2. Logistic regression (Model 1) results

#### 2.1 Univariable analysis (Model 1*_univariable_*)

**Table S1:**
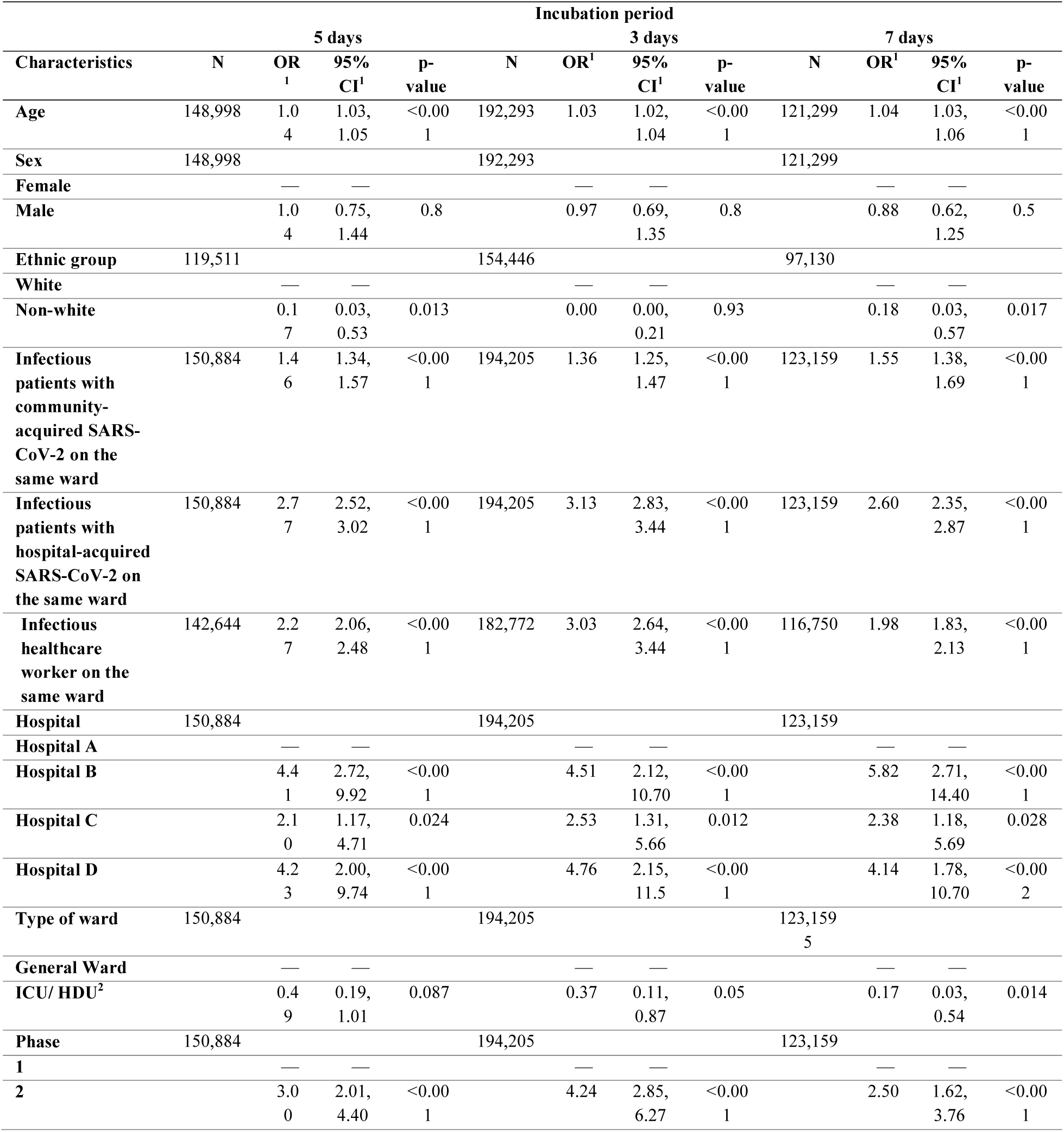
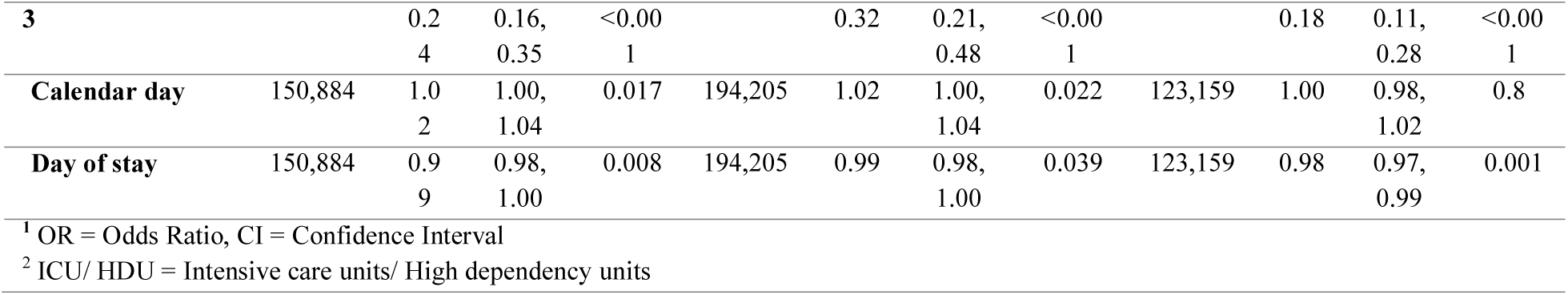
Univariable logistic regression results where the outcome is patient SARS-CoV-2 infection during the hospital stay (model *P*1*_univariable_*).

**Table S2:**
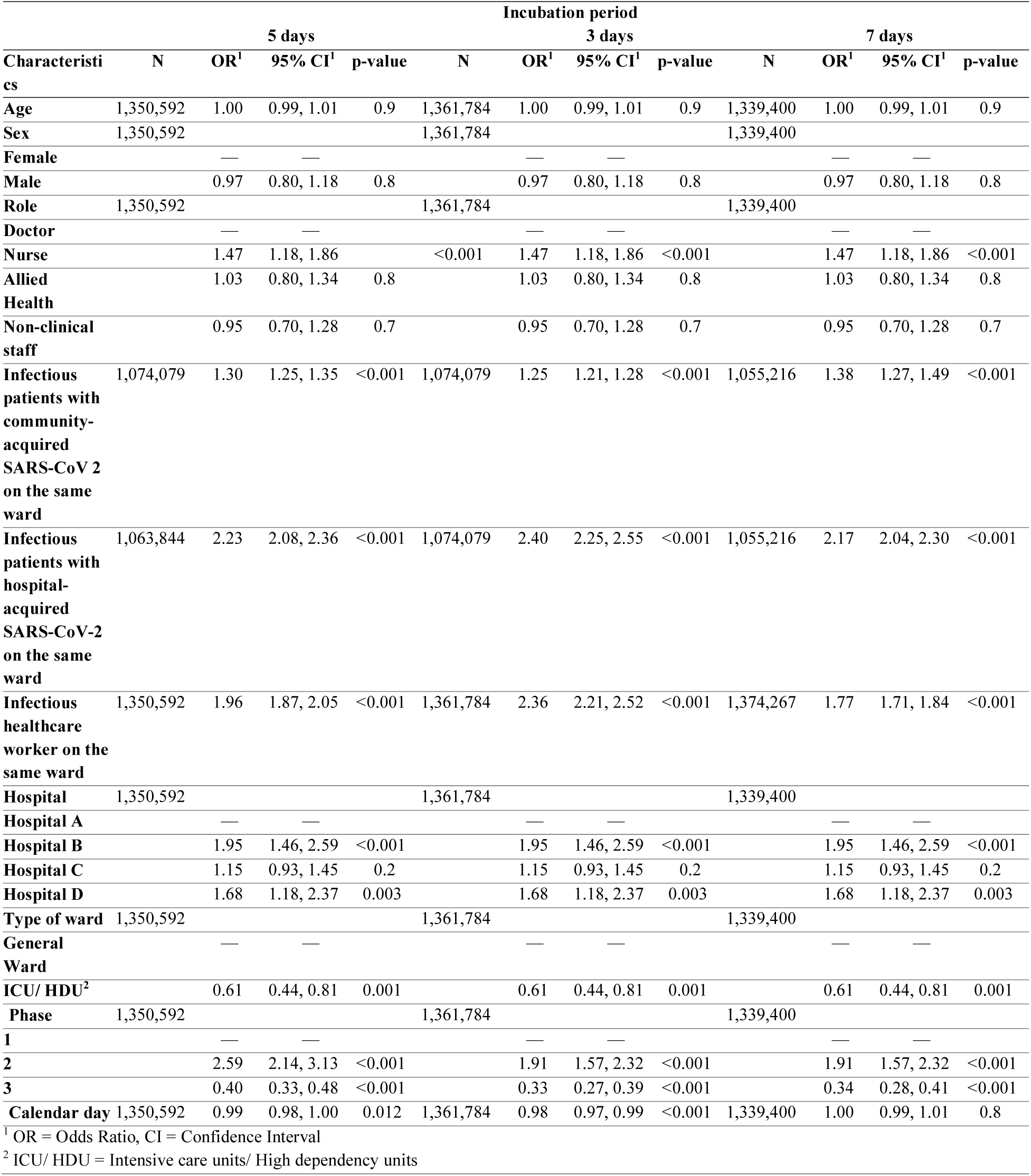
Univariable logistic regression results where the outcome is healthcare worker COVID-19 infection during the hospital stay (model *H*1*_univariate_*).

### 3. Generalised additive model (Model 2) results

#### 3.1 Daily risk of patient nosocomial SARS-CoV-2 infection (model *P*2)

**Figure S2.**
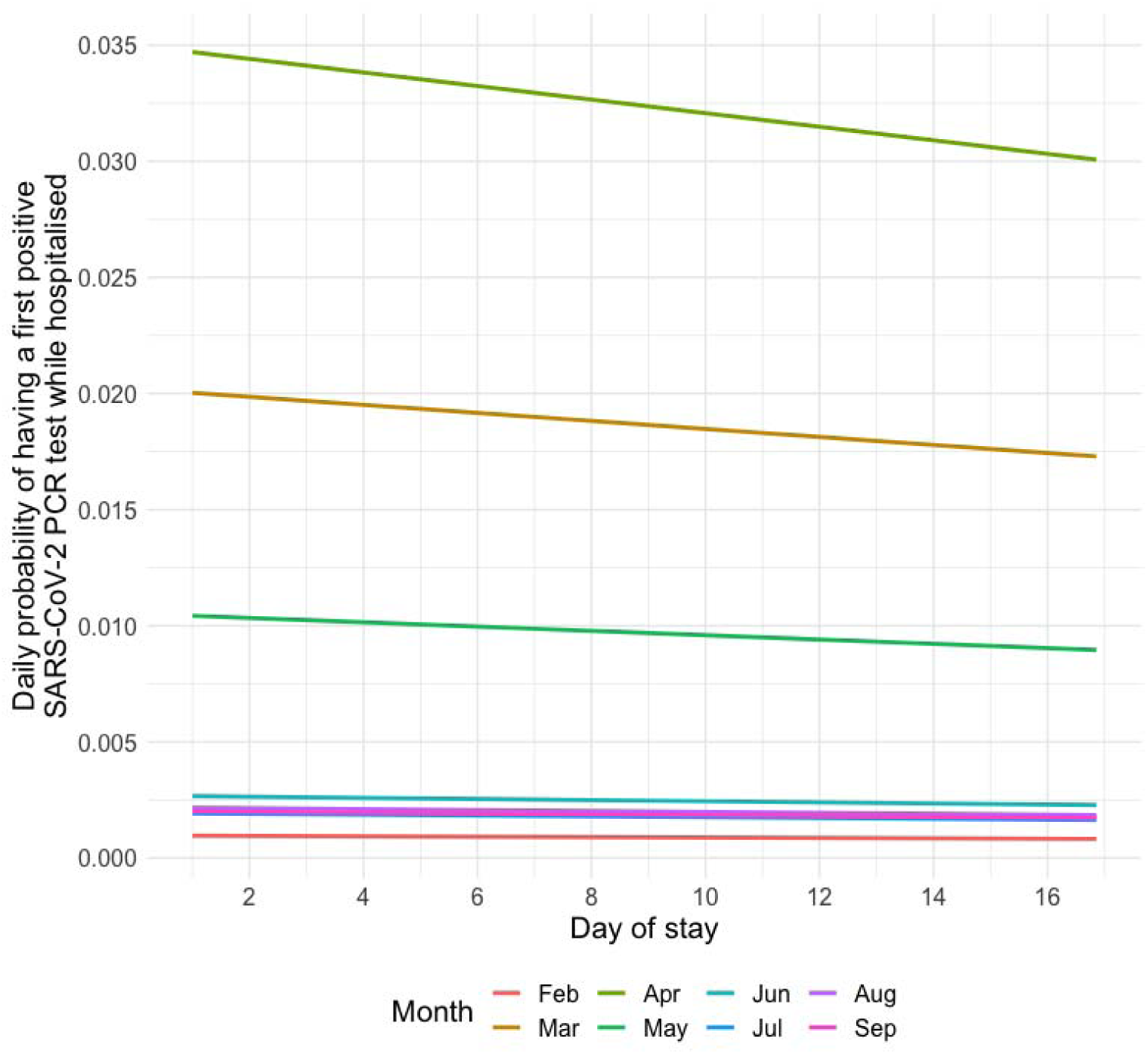
Daily probability of having a first positive SARS-CoV-2 PCR test during hospitalisation. The coloured lines represent the daily probabilities of having the first positive SARS-CoV-2 PCR test throughout a patient’s hospitalisation for months from February to September 2020. These probabilities are predictions from the generalised additive model with a logit link, with the binary outcome of assumed acquisition (yes/no) on each day as the dependent variable, and infectious patients and healthcare workers as the independent variables. Infectious patients were classified as having nosocomial SARS-CoV-2 infections with the assumption of a 5-day incubation period.

### 4. Generalised linear model with identity link (Model 3)

#### 4.1 Model comparison

To quantify the daily transmission risk posed by infectious patients and healthcare workers, we used a generalised linear mixed model with an identity link, thus allowing for the daily probability of infection to scale linearly with infection pressure from healthcare workers and patients and for their effects to be additive. Two models, one with interaction terms between the phases and forces of infection from patients and healthcare workers, and one without the interaction terms, were compared. Between these transmission models, the model with the best fit to data by WAIC was the one without interaction terms, which has an intercept (α), representing the infection risk not explained by covariates, and slopes (*beta*) which represent the infection risk associated with infectious patients (community- and hospital-acquired) and healthcare workers.

**Table S3:**
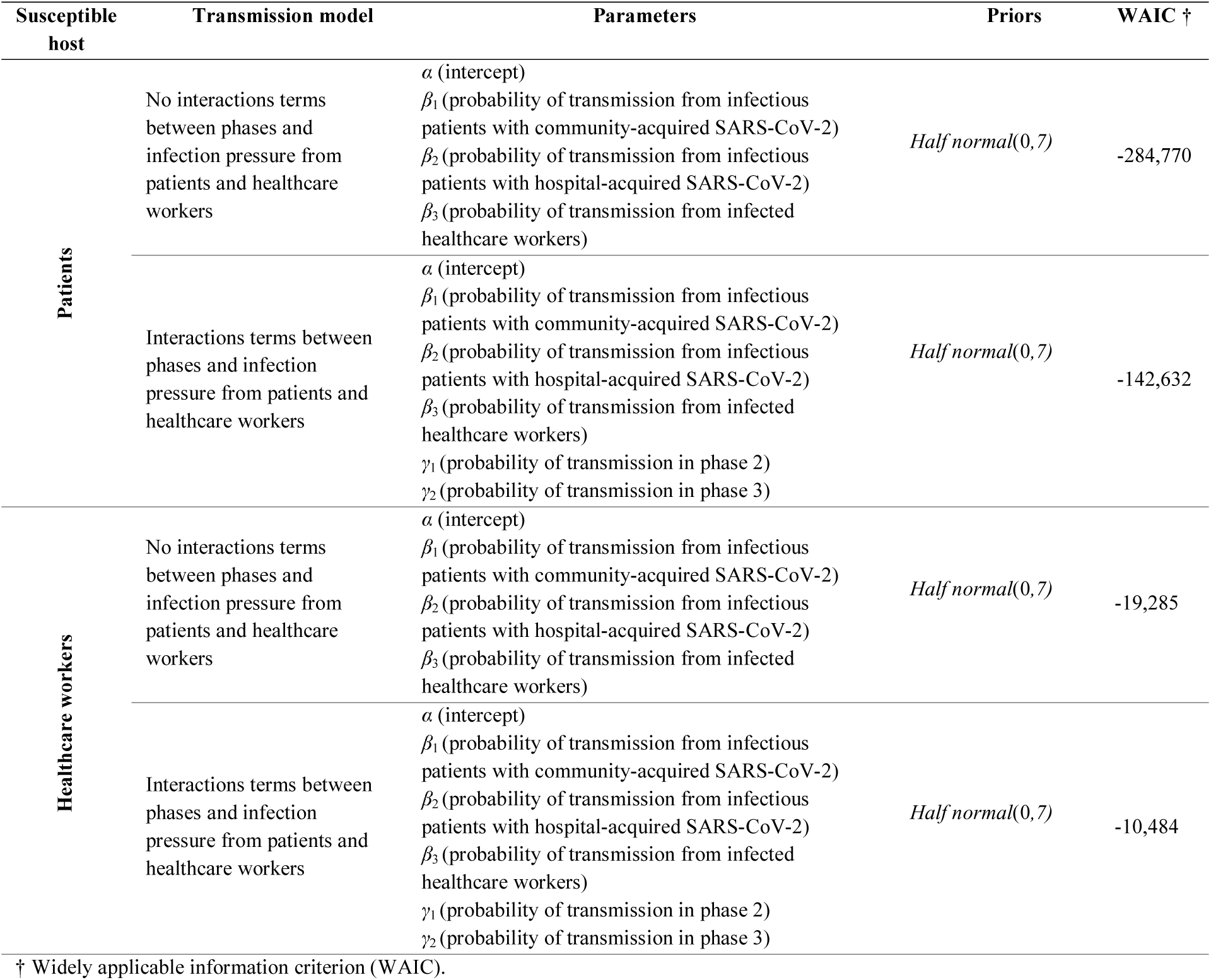
Comparison of widely applicable information criterion between a model with no interaction terms between phases and infection pressure from patients and healthcare workers versus a model with interaction terms.

#### 4.2 Model assessment of the main analysis models

Prior distributions were selected to be weakly informative half-normal distributions, such that the prior values are kept positive. We assessed the models using measures of Markov chain convergence including effective sample sizes and *R*^ which indicate if the chains had run for long enough and had mixed well.

Plots of iterations vs. sampled values for model parameters in the MCMC chains. The three different chains are plotted using different colours.

In the main analysis model where the outcome is hospital-acquired SARS-CoV2 infection amongst the patients, the *R*^ values were about 1 and the minimum effective sample size was 1700 across all parameters. The chains mixing is shown below.

**Figure S3.**
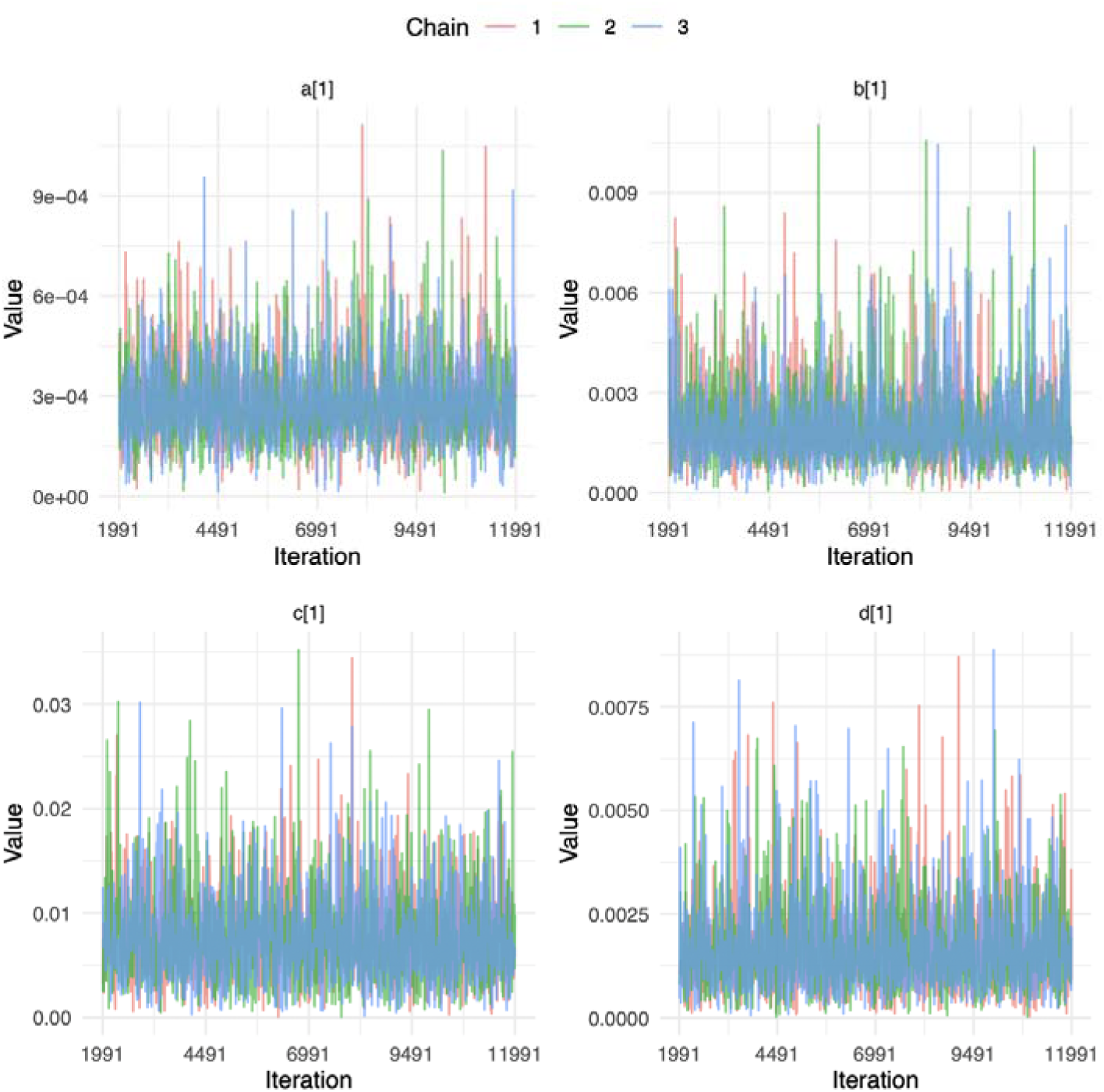
Model where outcome is patient SARS-CoV-2 infection acquired during hospitalisation. First plot of each parameter, representing a single ward is shown.

In the main analysis model where the outcome is hospital-acquired SARS-CoV2 infection amongst the HCW, the *R*^ values were about 1 and the minimum effective sample size was 1500 across all parameters. The chains mixing is shown below.

**Figure S5:**
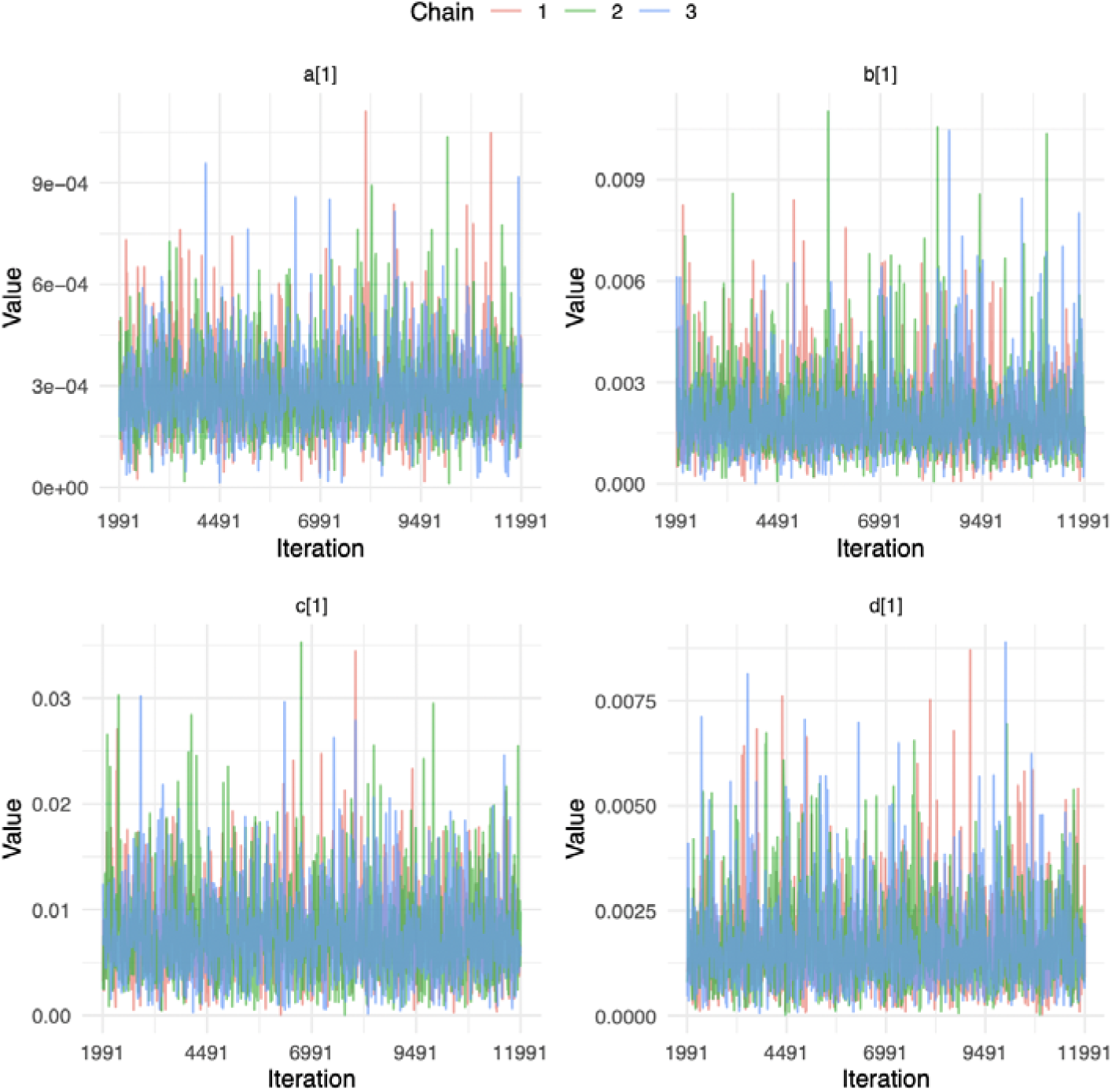
Model where outcome is healthcare worker SARS-CoV-2 infection. First plot of each parameter, representing a single ward is shown.

### 5. Sensitivity analysis

#### 5.1 Infectiousness scaled by day of infection according to incubation period

**Table S4:** The main analysis considers infectiousness to be binary, i.e., absolute numbers of infectious patients and healthcare workers in a ward on a particular day were used. Sensitivity analysis considered infectiousness to be scaled according to the time since the day of infection which, in turn, is based on an assumed incubation period of five days. This scaling of the number of infectious patients and healthcare workers in a ward on a particular day makes use of the relative infectiousness distribution derived by He *et al* [25] such that the sum of daily terms for a single infected patient who was present in the ward throughout their entire infectious period would equal one. Hence, the scaled parameters are an order of magnitude higher than the binary infectiousness model estimates.

| Infectious population posing transmission risk | Additional risk of acquiring nosocomial SARS-CoV-2 (%)<br>(mean, 95% credible interval) |  |  |  |
| --- | --- | --- | --- | --- |
|  | For susceptible patients |  | For susceptible healthcare workers |  |
|  | Binary infectiousness<br>(main analysis) | Scaled infectiousness | Binary infectiousness<br>(main analysis) | Scaled infectiousness |
| Background infection risk including undetected cases | 0.03 (0.02-0.03) | 0.03 (0.03-0.04) | 0.02 (0.02, 0.03) | 0.03 (0.02, 0.04) |
| Patients who acquired the infection from the community on the same ward | 0.20 (0.16-0.22) | 1.96 (1.6-2.23) | 0.05 (0.03, 0.07) | 0.60 (0.33, 0.76) |
| Patients who acquired the infection from the hospital on the same ward | 0.75 (0.55-0.95) | 6.56 (5.33-7.94) | 0.11 (0.08, 0.15) | 1.01 (0.70, 1.32) |
| Healthcare workers on the same ward | 0.17 (0.13-0.22) | 1.45 (1.01-1.91) | 0.10 (0.04, 0.20) | 0.83 (0.31, 1.62) |

#### 5.2 Sensitivity to choice of prior distributions

**Table S5:** Prior distributions for the various parameters were changed to test the sensitivity of the estimates to the choice of prior distributions.

| Infectious population posing transmission risk | Additional risk of acquiring nosocomial COVID-19 (%)<br>(mean, 95% credible interval) |  |  |  |
| --- | --- | --- | --- | --- |
|  | For susceptible patients |  | For susceptible healthcare workers |  |
| | $\alpha$: half normal(0, 7)<br>$\beta$: half normal(0, 7)<br>(main analysis) | $\alpha$: half normal(0, 7)<br>$\beta$: half normal(0.01, 3) | $\alpha$: half normal(0, 7)<br>$\beta$: half normal(0, 7)<br>(main analysis) | $\alpha$: half normal(0, 7)<br>$\beta$: half normal(0.01, 3) |
| Background infection risk including undetected cases | 0.03 (0.02-0.03) | 0.03 (0.02-0.03) | 0.02 (0.02, 0.03) | 0.02 (0.02, 0.03) |
| Patients who acquired the infection from the community on the same ward | 0.20 (0.16-0.22) | 0.20 (0.16-0.22) | 0.05 (0.03, 0.07) | 0.05 (0.03, 0.07) |
| Patients who acquired the infection from the hospital on the same ward | 0.75 (0.55-0.95) | 0.74 (0.56-0.95) | 0.11 (0.08, 0.15) | 0.11 (0.07, 0.15) |
| Healthcare workers on the same ward | 0.17 (0.13-0.22) | 0.16 (0.13-0.22) | 0.10 (0.04, 0.20) | 0.11 (0.04, 0.20) |

### 6. Model codes

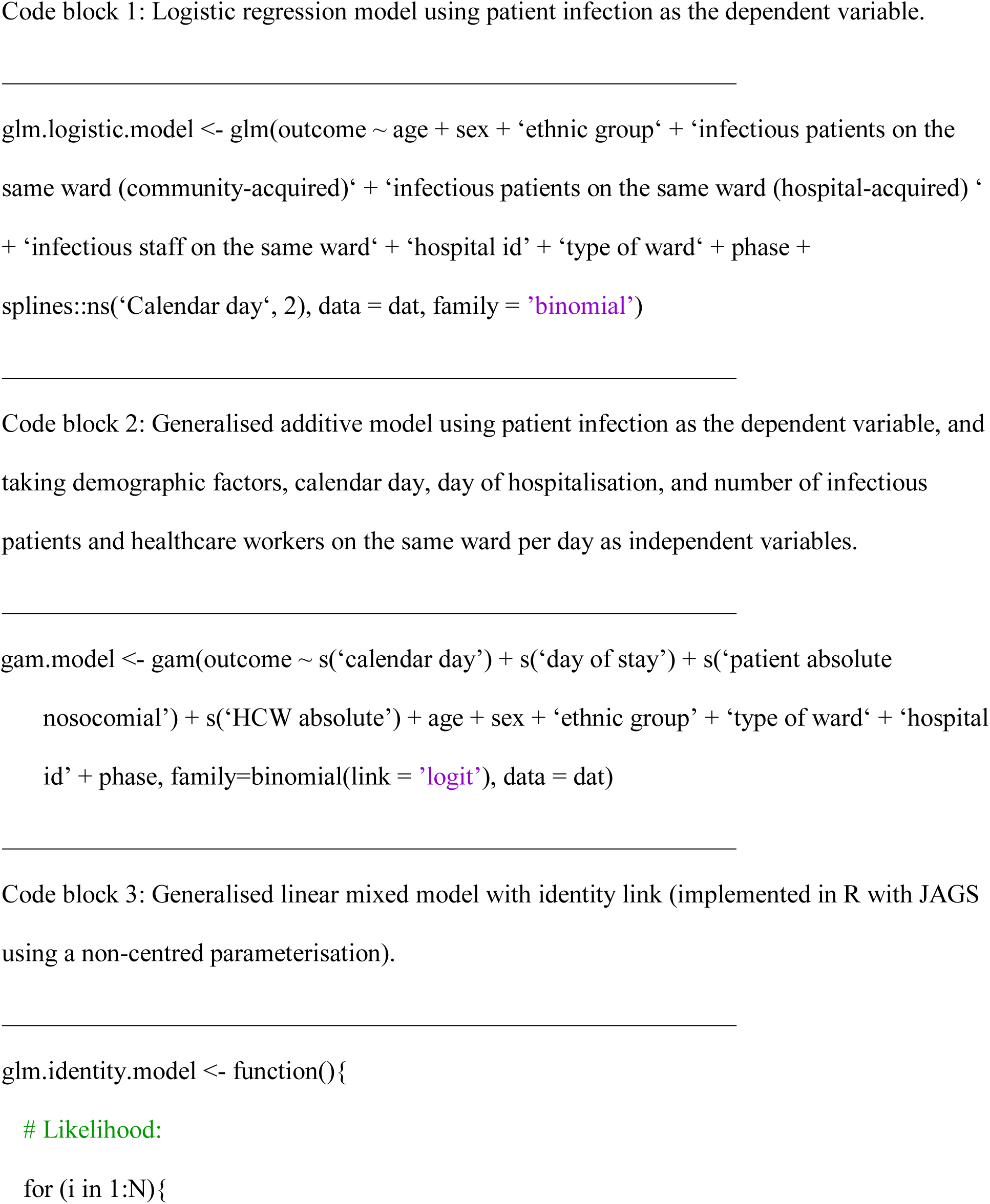
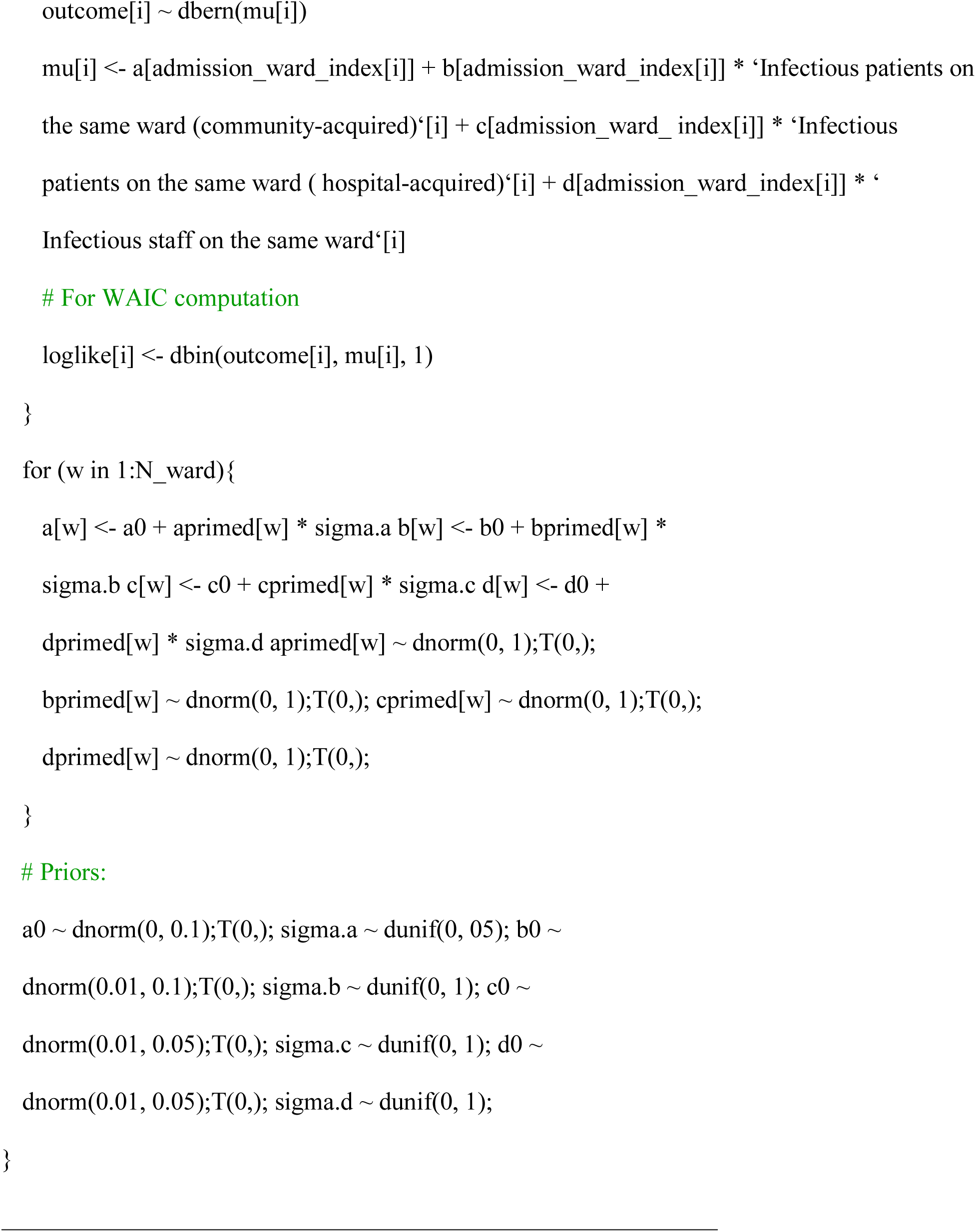

### 7. Combined nasal and oropharyngeal swabs

RT-PCR was performed using the Public Health England SARS-CoV-2 assay (targeting the RdRp gene), one of five commercial assays: Abbott RealTime (targeting RdRp and N genes; Abbott, Maidenhead, UK), Altona RealStar (targeting E and S genes; Altona Diagnostics, Liverpool, UK), Cepheid Xpert® Xpress SARS-CoV-2 (targeting N2 and E; Cepheid, California, USA), BioFire® Respiratory 2.1 (RP2.1) panel with SARS-CoV-2 (targeting ORF1ab and ORF8; Biofire diagnostics, Utah, USA), Thermo Fisher TaqPath assay (targeting S and N genes, and ORF1ab; Thermo Fisher, Abingdon, UK) or using the ABI 7500 platform (Thermo Fisher, Abingdon, UK) with the US Centers for Disease Control and Prevention Diagnostic Panel of two probes targeting the N gene.

### 8. Distributions of incubation period and generation time

**Figure S5:**
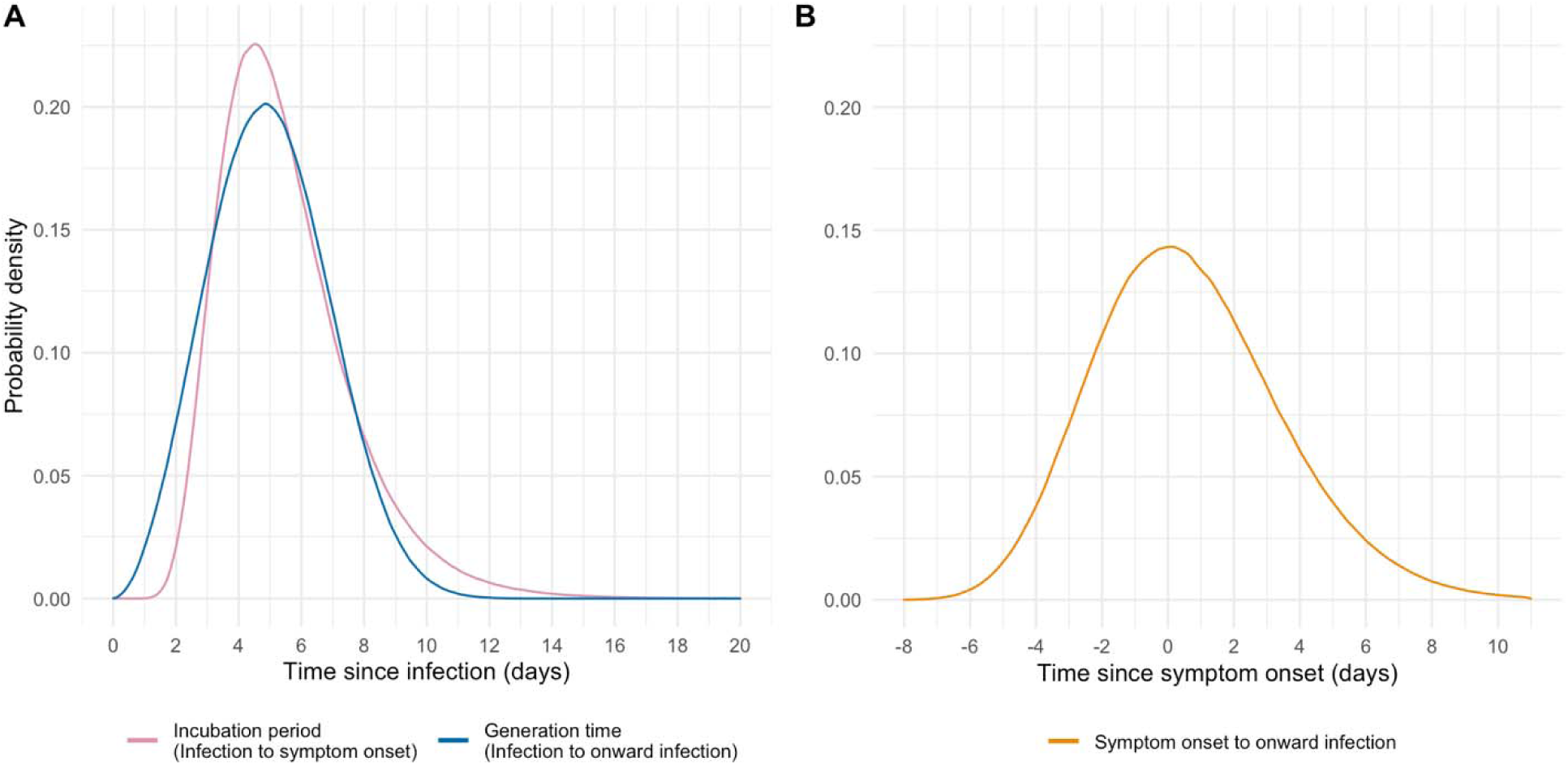
Distribution of the incubation periods, generation time (Panel A), and from symptom onset to onward infection (Panel B).

## 9. Oxford COVID infection review team

University of Oxford Medical School

- Hannah Chase
- Ishta Sharma
- Sarah Peters
- Archie Lodge
- Sai Parepalli
- Raghav Sudarshan
- Hannah Callaghan
- Imogen Vorley
- Gurleen Kaur
- Emel Yildirim
- Naomi Hudson

Oxford University Hospitals

- Omar Risk
- Tamsin Cargill
- Grace Barnes
- Josh Hamblin
- Jenny Tempest-Mitchell
- Ashley Elder
- Danica Fernandes
- Bara’a Elhag
- Edward David
- Rumbi Mutenga
- Dylan Riley

